# NeoGx: Machine Learning to Predict Genetic Evaluation Need in the Level IV NICU

**DOI:** 10.1101/2024.06.24.24309403

**Authors:** Austin A. Antoniou, David M. Gordon, Ashley Kubatko, Peter White, Bimal P. Chaudhari

## Abstract

**Objective:** Genetic disease is common in Level IV Neonatal Intensive Care Units (NICUs), yet clinicians often struggle to identify infants who would benefit from genetic evaluation. We developed and validated NeoGx, a machine learning (ML) algorithm using electronic health record (EHR) data to predict, early in the NICU stay, which neonates will require genetic evaluation within 18 months of life, enabling high-yield testing, including rapid genome sequencing (rGS), to be directed to the right infants while most beneficial.

**Methods:** Data were extracted from the EHRs of 14,272 Level IV NICU patients including structured data and phenotypes derived from clinical text. Patients were temporally divided into development (N=ll,201), calibration (N=1,080), and validation (N=1,991) cohorts. ML models were optimized using 3-fold cross validation in the development cohort to predict genetic evaluation by 18 months, then evaluated in an independent validation cohort.

**Results:** Using predictions accumulated over four NICU weeks, NeoGx achieved a ROC AUC of 0.849 and PR AUC of 0.771. NeoGx-guided referral reduced the mean time to first genetic evaluation from 44 to 29 days. When paired with rGS as the first-line test, the share of genetic cases reaching a definitive testing endpoint within 14 days rose from 9.5% to 68.6%.

**Conclusions:** NeoGx identifies Level IV NICU infants likely to need genetic evaluation early in their stay. Acting on its predictions could advance evaluation by an average of 15.2 days. When integrated with rGS, this approach can shorten the time to diagnosis, enabling timely management and improved outcomes for critically ill neonates.

**LAY SUMMARY:** Genetic conditions are a common cause of serious illness in newborns, but identifying affected infants early can be challenging. As a result, many families wait months or years for answers. We developed NeoGx, a machine learning tool that analyzes information already available in the electronic health record to identify NICU patients who may benefit from genetic evaluation within the first 18 months of life. Using data from more than 14,000 infants, we found that NeoGx could identify high-risk babies early in their hospitalization. Simulations suggest that combining NeoGx with rapid genomic sequencing could substantially shorten the time to genetic evaluation and diagnosis, helping families receive answers sooner and supporting earlier clinical decision-making.

## BACKGROUND AND SIGNIFICANCE

Genetic diseases are a leading cause of infant morbidity and mortality[1-4]. As such, they are a common cause of Neonatal Intensive Care Unit (NICU) admission, particularly to the highest acuity NICUs, designated Level IV by the American Academy of Pediatrics[1,5-7]. While common genetic diagnoses, such as classical aneuploidies (trisomies 13,18, and 21), are typically recognized quickly[2–4], the more than 10,000 rare genetic disorders often present with complex phenotypes not easily recognized by neonatologists or even by geneticists unfamiliar with the neonatal presentations [8,9]. This diagnostic challenge is particularly salient in Level IV NICUs, where high-acuity surgical needs and comorbidities related to prematurity can mask the subtle indicators of an underlying genetic condition[10]. Indeed, even though genetic disorders are relatively common in NICUs, many patients do not receive a diagnosis until after discharge[11,12]. In such a high acuity care setting, and in an era where precision therapies are increasingly available[13], there is urgent need to shorten the diagnostic odyssey for these neonates.

Genomic Sequencing (GS) is capable of making almost all genetic diagnoses for which an etiology is known[14]. Rapid GS (rGS) has been implemented in several patient care settings with turnaround time on the order of days rather than weeks [15]. The combination of comprehensiveness and timeliness has led providers and patient families alike to report high utility for rGS, even in cases where no diagnosis was made[16]. While rGS has a higher unit cost than narrower tests, the broader, more comprehensive nature of such testing makes it a more cost-effective choice, particularly early in a NICU course (within 14 days of ICU admission) [17,18]. However, these economic and clinical advantages are time-dependent; delays in recognizing an infant’s need for genetic evaluation often push the return of results past this critical window.

Much of the burden of making genetic testing decisions in the NICU lies with neonatology providers, who may not be equipped or confident to make these decisions[19,20]. It is at this stage that providers and patients could benefit from machine-aided predictions of who is likely to need genetic evaluation in the future. We developed NeoGx, a machine learning (ML) model to predict the receipt of early-in-life (prior to 18 months of age) genetic evaluation based on demographic and phenotypic data derived from the Electronic Health Record (EHR) during the first four weeks in the Level IV NICU.

Compared to previous efforts, we study a larger cohort of neonates, investigate more complex model architectures and feature sets, and predict a more inclusive target. Instead of predicting a specific molecular diagnosis or choice of a specific test such as microarray [21] or GS[22,23], we aim to predict which patients will eventually require any genetic evaluation. By functioning as a clinical trigger rather than a diagnostic endpoint, NeoGx is designed to accelerate the diagnostic process by augmenting testing practices in Level IV NICUs while remaining agnostic as to the current state of the art in critical care genetic testing.

## MATERIALS AND METHODS

Our study population consisted of 14,272 patients born between 1 January 2010 and 30 September 2024 and admitted to the Nationwide Children’s Hospital (NCH) Level IV NICU **(Supplementary Materials: Cohort and Data).** 124 subjects missing data for gestational age (GA) or birth weight (BW) were excluded. To naturally stratify with respect to demographics and clinical outcomes, as well as to create a validation cohort that most resembles present-day NICUs, the full cohort was temporally split by birthdate, a partitioning approach which has previously been effective and is regarded as preferable to a random partition of internal validation data[24,25]. The training cohort consisted of 11,201 patients born 2010-2021, after excluding 304 with classical aneuploidies and 149 who received genetic evaluation prior to Level IV NICU admission, to avoid biasing the model toward the easiest and most common neonatal diagnoses. A calibration cohort was made from the 1,080 patients born in 2022, and a validation cohort from the remaining 1,991 patients **(Figure 1).**

**Figure 1:**
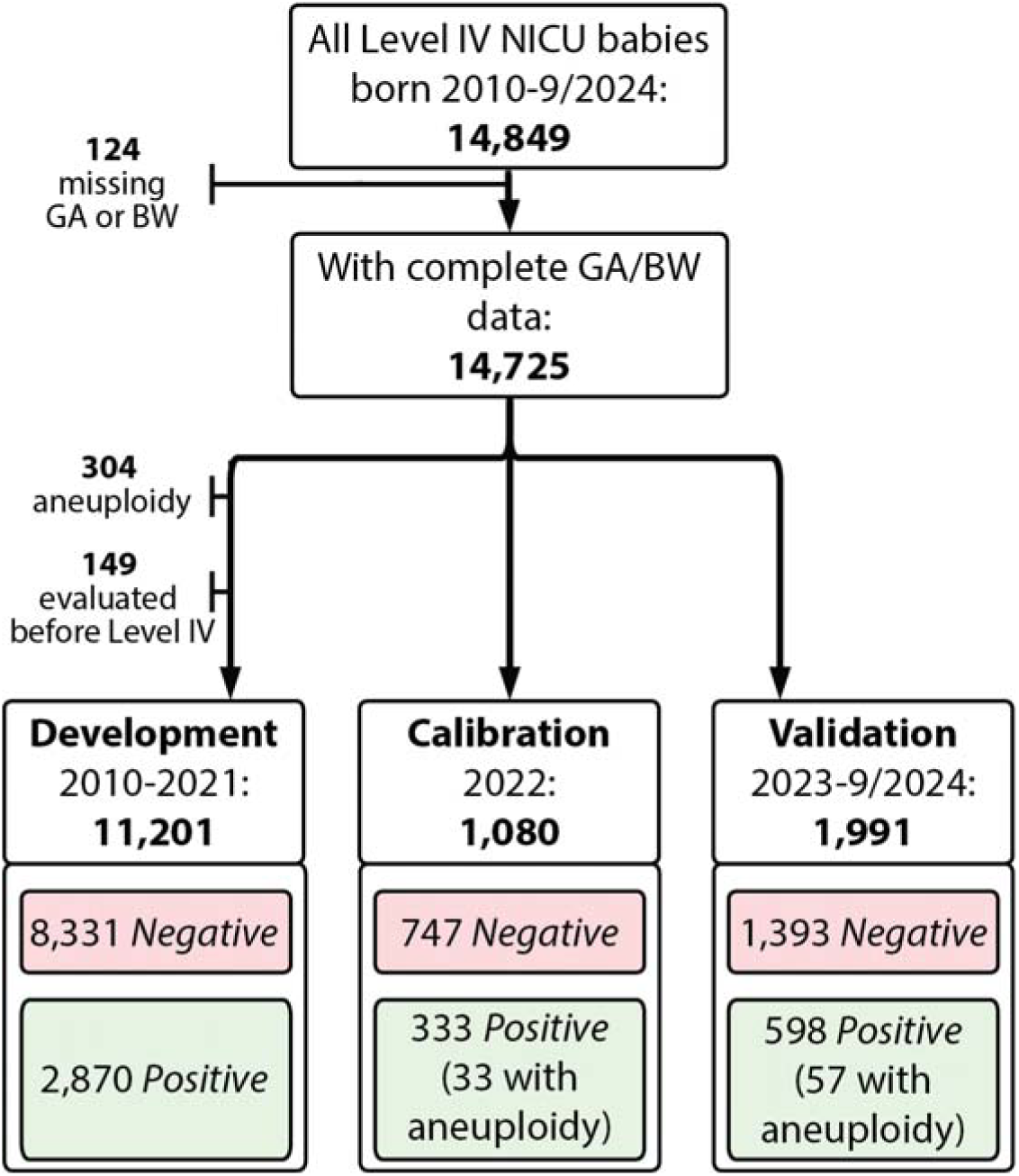
Study flow diagram for training, calibration, and validation cohorts. The full data set was temporally split by birth date to partition it into sub-cohorts: Development, for model selection and training; Calibration, for re-scaling classifier probability scores; and Validation, for calculating statistical performance metrics and estimating clinical impact. Each step in the diagram shows how many subjects met (and how many failed to meet) the inclusion criteria. Subjects missing gestational age (GA) or birthweight (BW) data were excluded. Subjects with classical aneuploidies were excluded from training set but were considered part of the positive class in the other cohorts.

Subjects were labelled *genetic* (the positive class) if they had a genetic laboratory test, diagnosis code, clinical consultation, or referral before 18 months of age **(Supplementary Materials: Curation and Table S1);** all others were labelled *non-genetic* (the negative class). The 18-month threshold was chosen under the rationale that genetic evaluation started after this age might have been motivated by common developmental and behavioral concerns rather than problems salient to their NICU hospitalization. In the entire cohort there were 3,801 genetic and 10,471 non-genetic subjects (Table 1).

**Table 1.** Summary of overall cohort composition and partitioned sub-cohorts. For categorical variables, the overall counts and percentage of each as well as within the development, calibration, and validation cohorts, respectively, are shown. For continuous variables, the mean and standard deviation are shown.0 Genetic evaluation by age 18 months was used as the label for machine learning classification training. Patients diagnosed with classical aneuploidies or who had genetic evaluation prior to Level IV NICU admission were excluded from the development cohort. (Amer.: American; Nat.: native; P.I.: Pacific Islander; wks: weeks; kgs: kilograms; SD: standard deviation; GA: gestational age; BW: birth weight; mo: months; Dx: diagnosis; GS: genomic sequencing).

|  |  | Overall | 2010-2021:<br>Development | 2022:<br>Calibration | 2023 -<br>2024(Q3):<br>Validation |
| --- | --- | --- | --- | --- | --- |
| Sex, n<br>(%) | Male | 8,017 (56.2) | 6,329 (56.5) | 607 (56.2) | 1,081 (54.3) |
|  | Female | 6,255 (43.8) | 4,872 (43.5) | 473 (43.8) | 910 (45.7) |
| Race, n<br>(%) | White | 9,769 (68.4) | 7,734 (69.0) | 718 (66.5) | 1,317 (66.1) |
|  | Black or African Amer. | 2,326 (16.3) | 1,776 (15.9) | 193 (17.9) | 357 (17.9) |
|  | Multiple | 1,061 (7.4) | 825 (7.3) | 79 (7.3) | 157 (7.9) |
|  | Unknown | 787 (5.5) | 618 (5.5) | 65 (6.0) | 104 (5.2) |
|  | Asian | 299 (2.1) | 228 (2.0) | 22 (2.0) | 49 (2.5) |
|  | Amer. Indian Or Alaska Nat. | 23 (0.2) | 15 (0.1) | 3 (0.3) | 5 (0.3) |
|  | Nat. Hawaiian or Other P.I. | 7 (0.0) | 5 (0.0) | 0 (0.0) | 2 (0.1) |
| Ethnicity,<br>n (%) | Not Hispanic or Latino | 13,317 (93.3) | 10,461 (93.4) | 1,017 (94.2) | 1,839 (92.4) |
|  | Other/Unknown | 687 (4.8) | 557 (5.0) | 42 (3.9) | 88 (4.4) |
|  | Hispanic or Latino | 268 (1.9) | 183 (1.6) | 21 (1.9) | 64 (3.2) |
| Gestational Age (weeks), mean (SD) |  | 34.8 (5.0) | 34.8 (5.0) | 34.8 (5.0) | 34.7 (4.9) |
| Birthweight Z-Score (Olsen 2010), mean (SD) |  | -0.1 (1.3) | -0.1 (1.3) | -0.2 (1.4) | 0.0 (1.2) |
| Genetic Evaluation Start ≤ 18mo, n (%) |  | 3,801 (26.6) | 2,870 (25.6) | 333 (30.8) | 598 (30.0) |
| Genetic Diagnosis ≤ 18mo, n (%) |  | 674 (4.7) | 448 (4.0) | 78 (7.2) | 148 (7.4) |
| Classical Aneuploidy Diagnosis, n (%) |  | 90 (0.6) | 0 (0.0) | 33 (3.1) | 57 (2.9) |
| Had Genomic Sequencing, n (%) |  | 659 (4.6) | 370 (3.3) | 84 (7.8) | 205 (10.3) |
| Total, n |  | 14,272 | 11,201 | 1,080 | 1,991 |

### Model Development

Models incorporated baseline static features, including age at admission, sex, GA, and BW Z-scores, alongside longitudinal phenotypic profiles updated daily to capture the evolving clinical presentation over the first four weeks of Level IV NICU hospitalization **(Supplementary Materials - Model Development).** To represent EHR data, we compared two vocabularies: PhecodeX-mapped ICD codes and Human Phenotype Ontology (HPO) terms extracted from clinical text via natural language processing. Overlapping GA and BW terms were removed from both vocabularies to prevent feature correlation **(Supplementary Methods: Phenotype Features).**

A layered search strategy utilizing 3-fold cross-validation on Day 1 data was deployed to optimize feature encodings and classifier hyperparameters **(Supplementary Methods: Classifier and Hyperparameters),** using the mean Precision-Recall Area Under the Curve (PR-AUC) as the primary evaluation metric. To evaluate the impact of vocabulary choice and potential bias regarding prematurity, we analyzed four model variants across combinations of phenotype vocabulary (HPO vs. Phecodes) and GA inclusion (+/- GA). Final variants were retrained on the entire training cohort using Day 1 data and sigmoid calibrated **(Supplementary Methods: Calibration and Thresholding and Figure S1).** The HPO-based model without GA was ultimately selected to estimate potential clinical impact.

### Measuring classifier performance

Models were evaluated using Precision-Recall (PR) and Receiver Operating Characteristic (ROC) curves, with performance tracked across NICU days 1-7,14, 21, and 28. To ensure clinical feasibility, we set a probability threshold at the 70th percentile (score: 0.487), targeting a 30% positive prediction rate **(Supplementary Materials - Calibration and Thresholding).** Feature importance was assessed using SHapley Additive explanations (SHAP), with Spearman correlations used to determine the directionality of feature influence[26].

Beyond overall performance, we calculated precision, recall, and prediction rates for specific clinical subgroups (e.g., classical aneuploidies, pre-admission genetic evaluation, and patients who received GS). To identify potential algorithmic bias, we compared target class prevalence and model metrics across demographic subgroups (sex, race, ethnicity, and GA; **Supplementary Methods: Bias Evaluation).**

### Estimating clinical cost and benefit

To assess potential impact of implementing NeoGx as a clinical decision support tool, we simulated interventions that (i) accelerated first genetic evaluation to the day a patient received a positive NeoGx prediction or (ii) shortened evaluation duration by using only rGS as a first-line test. We also considered combinations of (i) and (ii). We calculated time to diagnosis or GS and non-GS tests avoided, balanced against increased intensity of test utilization under various simulated scenarios. **(Supplementary Materials - Estimated clinical cost and benefit).**

## RESULTS

We evaluated multiple approaches to phenotype representation, systematically varying the vocabulary (Phecode vs. HPO), feature selection, and encoding strategies **(Supplementary Materials: Phenotype features and Table S2).** Integrating these variations with diverse classifier architectures and hyperparameter configurations, we utilized a layered search to identify top-performing models based on 3-fold mean PR-AUC **(Supplementary Materials: Classifier and hyperparameters).** The initial candidates were both XGBoost based: one utilizing 2,419 binary Phecode features and the other 2,535 HPO terms weighted by bidirectional information content[27] **(Supplementary Materials: Selected Parameters).** Both models incorporated four static baseline features: age at admission, sex, BW Z-score, and GA.

### Separation and Classification Metrics Over Four ICU Weeks

Out of concern for bias against infants born premature, we evaluated four primary model variants: The two initial candidates, each with and without GA as a model feature, over the first 28 days of NICU admission[28] **(Figure 2a-b).** While on day 1, the Phecode model with GA showed strongest initial performance, the HPO-based models demonstrated rapid performance gains as clinical documentation accrued. Although the HPO model without GA initially performed worse on day 1 than the PheCode with GA model (ROC AUC 0.812; PR AUC 0.724 vs. ROC AUC 0.836; PR AUC 0.767], its ROC AUC surpassed both Phecode-based models by day 2, and its PR AUC surpassed their Phecode counterparts by day 5. Furthermore, adding GA to the HPO models had a negligible impact, with AUC scores for GA and non-GA versions remaining within 0.002 of each other through day 28. These relative performance trajectories remained consistent when restricting the analysis to patients remaining admitted on each respective prediction day **(Figure 2c-d],**

**Figure 2.**
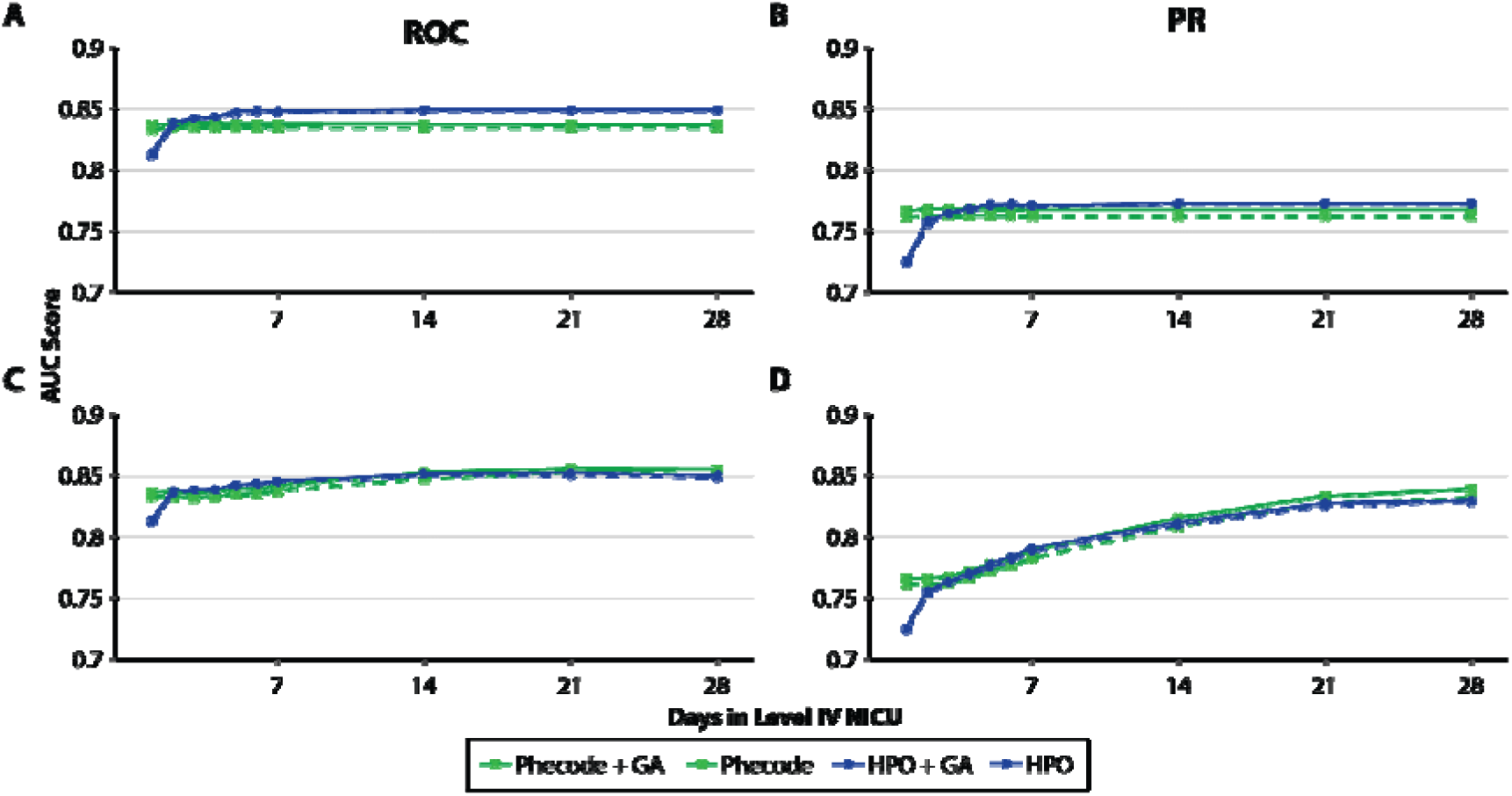
Performance for four classifier variants over 4 NICU weeks. Phecode-based models (green] and HPO-based models (blue] are shown with (solid lines] and without (dotted lines] gestational age (GA] as a feature, **(a]** Receiver Operating Characteristic (ROC] Area Under the Curve (AUC] and **(b]** Precision-Recall (PR] AUC scores over the 28-day period for the full validation cohort. To account for varying lengths of stay, **(c]** ROC AUC and **(d]** PR AUC scores are also presented for a restricted cohort consisting only of patients still admitted on each prediction day (censoring for death or discharge].

The HPO models were ultimately favored due to their distinct responsiveness to real-time phenotypic data accrual. Specifically, the HPO model without GA demonstrated superior recall for extremely premature infants (GA < 28 weeks), with negligible trade-offs in precision or overall performance across other GA strata **(Supplementary Materials - Bias Evaluation, Figure S2).** Consequently, the HPO-without-GA model was selected and is hereafter designated **NeoGx.** Based on the full validation cohort, using each patient’s maximum classifier score prior to discharge or Level IV NICU day 28, NeoGx achieved a ROC AUC of 0.849 (95% CI: 0.830 - 0.866) and a PR AUC of 0.771 (95% CI: 0.742 - 0.798) **(Figure 3a-b).** To simulate real-world clinical implementation, subsequent results are reported at a fixed probability threshold of 0.487 (the 70^th^ percentile of calibration cohort scores, selected to yield a clinically manageable alert volume of approximately 30%). At this threshold, NeoGx demonstrated a precision of 0.664 and a recall of 0.687 **(Figure 3c-d),** flagging 619 of the 1,991 validation patients (31.1%) as candidates for genetic evaluation.

**Figure 3.**
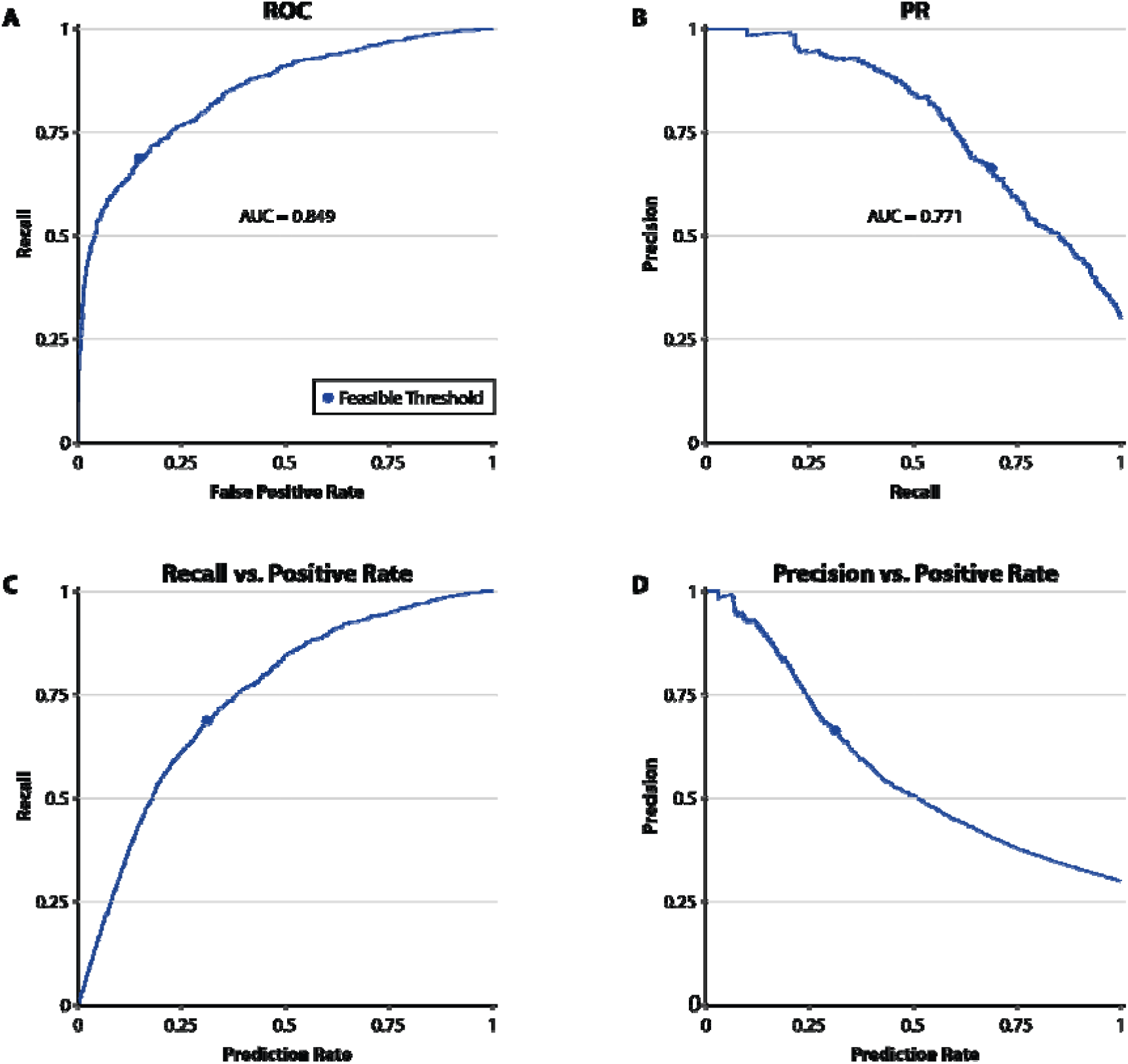
NeoGx performance metrics and clinical implementation curves. Separation and classification results are shown for the HPO-based model without gestational age **(NeoGx].** Each patient in the validation cohort was assigned the maximum predicted score recorded prior to discharge or day 28. Note: This "maximum score" logic captures the earliest point at which clinical evidence reached the decision threshold; most positive predictions (70.9%] were triggered on NICU day 1. Curves are parameterized by prediction probability threshold; the dot indicates performance at the selected clinical threshold (0.487], derived from the 70th percentile of scores in the calibration cohort, **(a] Receiver Operating Characteristic (ROC) curve** illustrating NeoGx’s ability to discriminate between infants who did and did not receive genetic evaluation, **(b) Precision-Recall (PR) curve** showing model performance in the context of the study’s 30% prevalence rate. **(c) Recall versus Positive Prediction Rate:** Illustrates the proportion of all genetic cases captured (Recall) relative to the percentage of the total NICU population flagged as positive (Prediction Rate). **(d) Precision versus Positive Prediction Rate:** Shows the proportion of flagged infants who truly received genetic evaluation (Precision) at varying population screening levels.

While our primary endpoint (any genetic evaluation within 18 months) is broader than typical literature targets, evaluating NeoGx requires understanding performance on other endpoints of interest which NeoGx is not optimized for. Of 411 infants in the positive class and flagged by NeoGx, 365 (88.8%) underwent genetic testing, and 122 of those tested (33.4%) received a genetic diagnosis. We evaluated NeoGx’s recall across five clinically meaningful subgroups within the positive class **(Table 2):** patients whose evaluation began before admission or post-discharge (representing the earliest and latest intervention windows), patients who received any genetic diagnosis, patients with a classical aneuploidy, and patients who underwent GS.

**Table 2.** Summary of genetic outcome subgroups of interest and the ML model’s recall of each. The first two groups are patients whose genetic evaluation began before level IV admission and after discharge, respectively. Evaluation start is defined by age of first genetic lab test, consult, referral, or diagnosis code. The next two groups are defined by having any genetic diagnosis code ("received Dx”) and by having a diagnosis code for a classical aneuploidy ("Classical Aneuploidy Dx”). The final group is made up of patients who, at some point in their genetic evaluation, received a GS test. (Abbreviations: "Eval”: "Evaluation”; "Dx”: "Diagnosis”; "GS”; "Genomic Sequencing”).

| Outcome subgroup | # Patients<br>(% positive class) | # Predicted positive<br>(% of subgroup) |
| --- | --- | --- |
| Eval. start < Level IV Admission | 104 (17.4%) | 80 (76.9%) |
| Eval. start > discharge | 100 (16.7%) | 36 (36.0%) |
| Received Dx | 151 (25.3%) | 122 (80.8%) |
| Classical Aneuploidy Dx | 57 (9.5%) | 51 (89.5%) |
| Received GS | 205 (34.3%) | 163 (79.5%) |

### Feature Importance

From SHAP scores, the phenotypes which most influenced positive predictions included abnormalities of the skeletal, cardiovascular, and genitourinary systems **(Table 3, Figure S3).** Top features influencing negative predictions included admission age, BW Z-score, and the presence of constitutional symptoms. Findings for other models are in **(Supplementary Materials: Feature Importance, Table S3).**

**Table 3.** Feature importance and directionality. Top 15 features ranked by their mean absolute SHAP value in the validation cohort. Mean SHAP values represent the average contribution of each feature to the model output. Spearman correlations between feature values and SHAP values were used to infer directionality. A positive correlation ("+") indicates that increasing feature values tended to increase model predictions, where as a negative correlation ("-") indicates that increasing feature values tended to decrease model predictions.

| Feature Name | Mean<br>Absolute<br>SHAP<br>Value | Mean<br>SHAP<br>Value | SHAP-Feature<br>Spearman<br>Correlation | Inferred<br>Direction |
| --- | --- | --- | --- | --- |
| Level IV Admit Age (days) | 0.21 | -0.04 | -0.85 | - |
| Abnormality of the musculoskeletal system (HP:0033127) | 0.16 | 0.04 | 0.87 | + |
| Abnormal heart morphology (HP:0001627) | 0.14 | 0.03 | 0.84 | + |
| BW Z-Score | 0.12 | -0.01 | -0.93 | - |
| Abnormal skeletal morphology (HP:0011842) | 0.09 | 0.02 | 0.66 | + |
| Constitutional symptom (HP:0025142) | 0.08 | 0.00 | -0.84 | - |
| Abnormality of head or neck (HP:0000152) | 0.08 | 0.01 | 0.87 | + |
| Abnormal cardiovascular system morphology (HP:0030680) | 0.08 | 0.01 | 0.83 | + |
| Abnormality of the genitourinary system (HP:0000119) | 0.07 | 0.01 | 0.82 | + |
| Abnormality of the skeletal system (HP:0000924) | 0.06 | 0.01 | 0.19 | + |
| Abnormal cerebral ventricle morphology (HP:0002118) | 0.05 | 0.00 | 0.56 | + |
| Abnormal renal morphology (HP:0012210) | 0.05 | 0.01 | 0.53 | + |
| Abnormality of the digestive system (HP:0025031) | 0.04 | 0.02 | 0.68 | + |
| Abnormal fetal morphology (HP:0034058) | 0.04 | 0.01 | 0.60 | + |
| Abnormality of limbs (HP:0040064) | 0.04 | 0.01 | 0.60 | + |

### Class and Model Bias

We examined bias both in the positive class label and in model performance. There was no apparent difference in positive class prevalence by sex, race, or ethnicity. There was a lower rate of genetic utilization among babies born at GA < 35 weeks, but this effect was mitigated by the choice of 18-month follow-up window **(Supplementary Materials - Bias Evaluation).** Similarly, we found no evidence of performance disparities across sex, race, or ethnicity subgroups. Performance differences were observed by GA, with reduced positive prediction rates, precision, and recall among extremely premature infants (<28 weeks GA) relative to infants born at ≥35 weeks GA. Notably, these differences were smaller in NeoGx than in alternate models **(Supplementary Materials - Bias Evaluation** and **Supplementary File - Bias Tests).**

### Projected Clinical Impact

Having established that NeoGx recalls the clinically relevant subgroups **(Table 2),** weights face-valid features **(Table 3),** and shows no significant bias by sex, race, or ethnicity, we next estimated the clinical impact of acting on its predictions. We simulated implementation strategies that used ML predictions to accelerate the initiation of genetic evaluation and, separately, the use of rapid genomic sequencing (rGS) to shorten the duration of the diagnostic workup. Under the ML-assisted initiation strategy, the mean age at first genetic evaluation decreased from 44 days to 29 days, corresponding to a mean reduction of 15.2 days (95% CI 11.0 - 19.5) **(Figure 4a).** The impact was greatest among extremely premature infants (GA < 28 weeks), for whom the mean reduction was 33.4 days (95% CI 18.8 - 51.8) **(Figure S4).** Those with latest genetic evaluation would receive the greatest benefit from NeoGx predictions: the 75^th^ percentile first genetic evaluation age would decrease from 31 days to 6 and the 90^th^ percentile from 133 days to 74.

**Figure 4.**
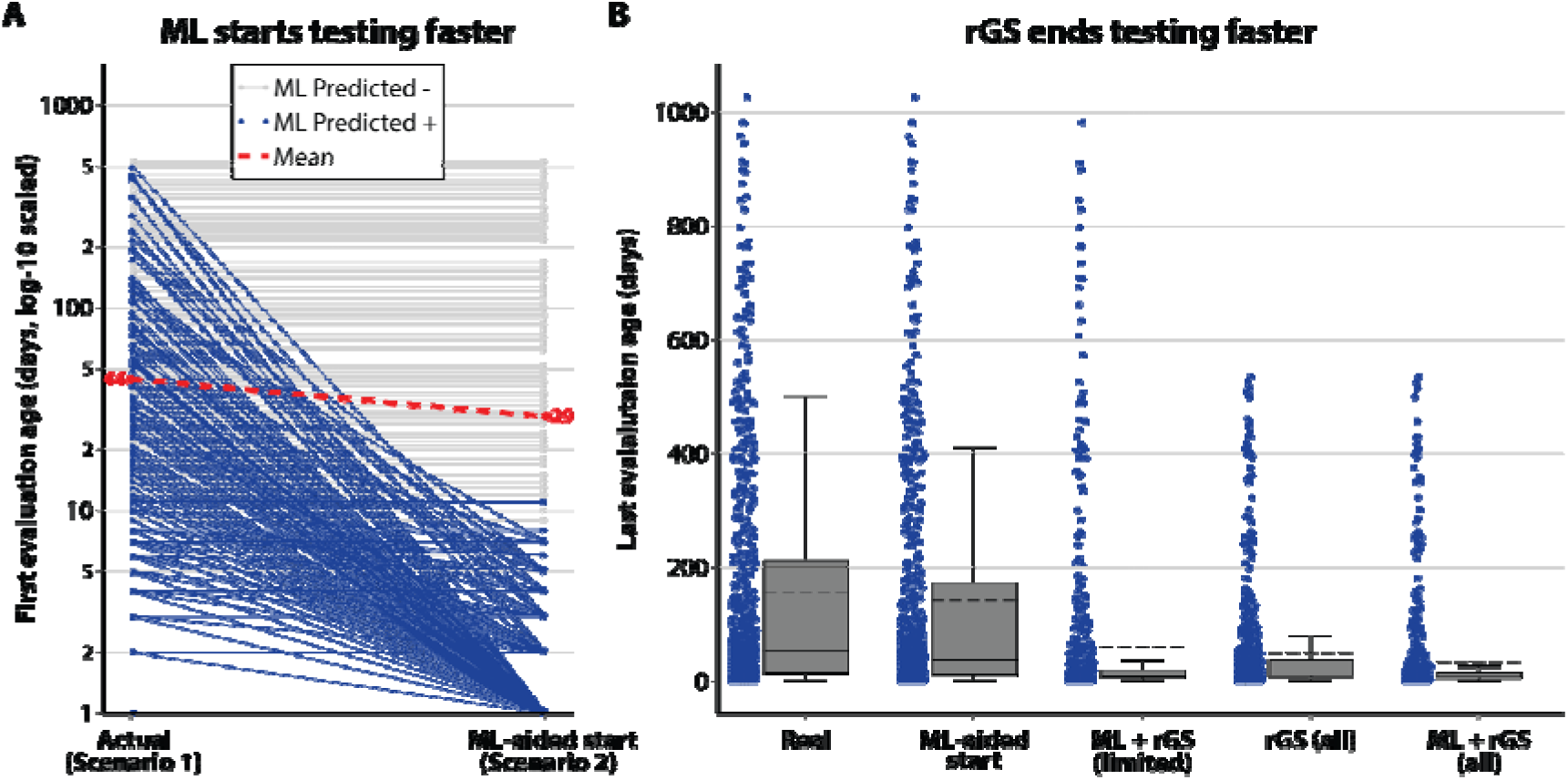
Simulated scenarios implementing intervention of ML to improve genetic evaluation start time and rGS to shorten the interval between first and last genetic evaluation event, (a]. Parallel coordinates comparison of actual age at first genetic evaluation versus simulated ML-assisted age at first genetic evaluation. Everyone who had received genetic evaluation is represented by a line, connecting their actual evaluation start age (left] to their simulated ML-assisted evaluation start age (right]. The vertical axis is measured in days, log-10 scaled. Individuals shown in blue had their time improved by ML due to a positive prediction; those shown in grey received a negative ML prediction and are assumed to receive zero benefit to genetic evaluation age. **(b]** Box plot showing the distribution of age at last genetic evaluation event, plus simulated time at last event under implementation scenarios 1-5 (See Methods - Estimating clinical cost and benefit].

We next evaluated the combined effects of earlier evaluation and rGS testing on the time required to complete genetic evaluation **(Figure 4b).** ML-assisted referral alone (Scenario 2) produced only a modest increase in the proportion of infants reaching a genetic diagnosis or completed GS by 14 days of age, from 9.5% under observed care (Scenario 1) to 11.9%. However, when rGS was assumed to be ordered at the time of an ML recommendation (Scenario 3), this proportion increased to 68.6%. In a hypothetical setting where rGS was used as the sole first-line genetic test for ah patients (Scenario 4), ML-assisted rGS ordering further increased the proportion of infants completing genetic evaluation by 14 days from 55.2% (Scenario 4) to 75.9% (Scenario 5) **(Figure S5).**

We also examined the patient-level implications of ML-assisted rGS (Scenario 3]. Under this scenario, NeoGx recommended rGS for 619 of 1,991 validation patients (31.1%], of whom 411 (66.4%] went on to receive a genetic evaluation in real-world care. Among these 411 true positives, 163 received GS during their clinical course at a mean age of 14.2 days; for these patients, acting on NeoGx’s recommendation would have advanced rGS initiation to a mean age of 1.2 days. A further 201 received only non-GS molecular or biochemical testing — workups an rGS-first strategy would replace with a single, higher-yield test — and 47 had only a clinical-genetics consult or referral on record with no testing performed; they and the remaining false positive 208 NeoGx recommendations did not align with a documented genetic test within 18 months and would have received rGS with an unknown outcome **(Figure 5).**

**Figure 5.**
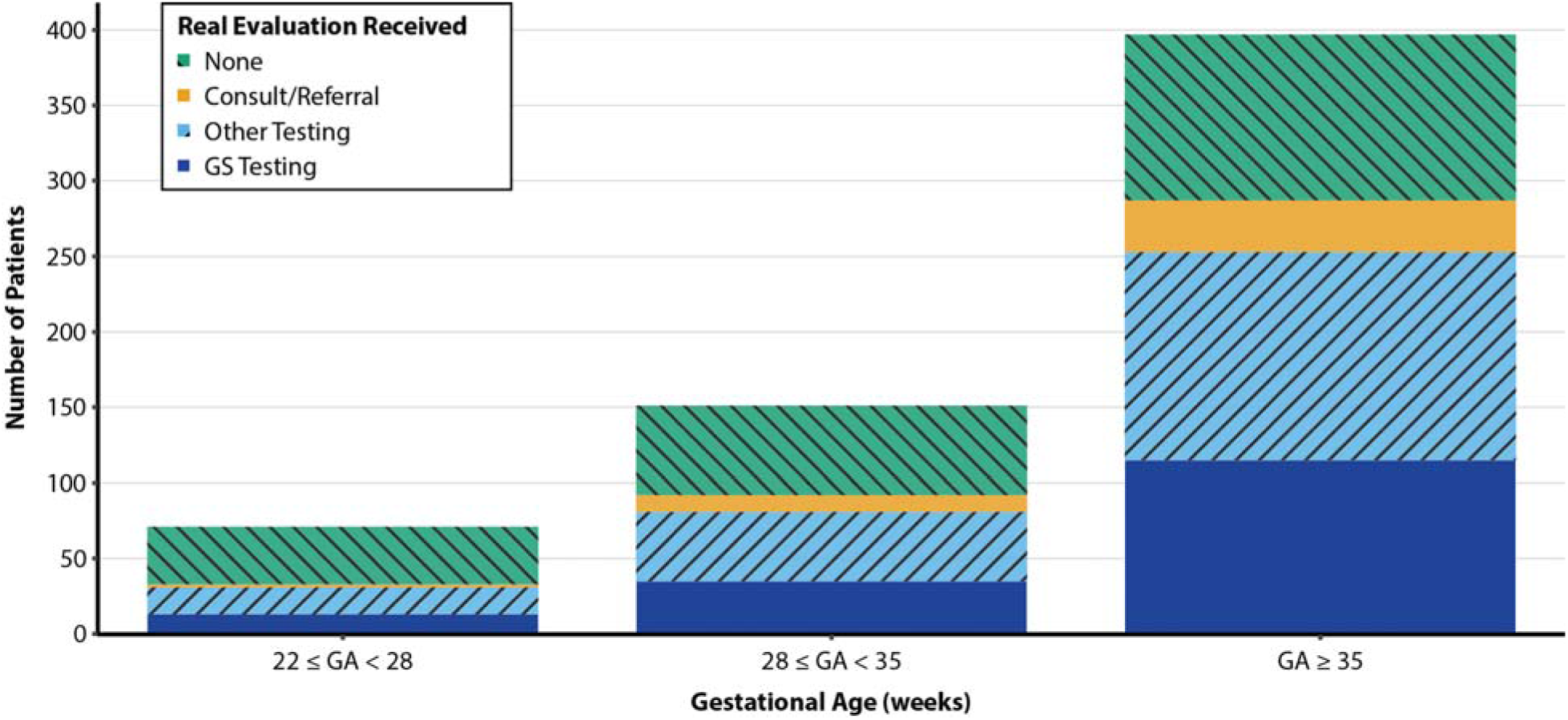
Patients recommended for genetic evaluation by NeoGx in simulation, by gestational age and by highest level of genetic evaluation actually received. All 619 patients with a positive NeoGx prediction are accounted for. Counts are grouped by GA category: 22 ≤ GA < 28, 28 ≤ GA < 35, and babies born at GA ≥ 35 weeks. Those who received Genomic Sequencing (GS) are indicated in dark blue; those who received non-GS molecular testing or biochemical testing are indicated in light blue; those who received no testing but had a consult or referral to genetics are indicated in orange. The remainder, in teal, received no genetic evaluation. Levels of evaluation are tiered, with GS being the highest tier, followed by other testing, followed by consult/referral. Patients who had evaluation events at one level may have one or more evaluation events at the lower levels.

Accounting for all non-GS molecular testing, NeoGx-flagged patients underwent 1,063 genetic tests that could have been bypassed by using rGS as a first-line diagnostic. In contrast, only 208 NeoGx-prompted rGS tests would be ordered for patients who did not otherwise receive any form of genetic evaluation within 18 months. Collectively, these dynamics demonstrate that an ML-assisted strategy could be used to motivate clinicians to redirect a substantial burden of longitudinal, stepwise testing towards earlier, more definitive testing.

## DISCUSSION

This study shows that routinely available EHR data can support early, phenotype-driven triage for genetic evaluation in a Level IV NICU. Rather than waiting for diagnostic suspicion to emerge through standard care, NeoGx enables a proactive referral model in which infants most likely to benefit from genetic testing can be identified near the start of admission. The modeled benefit among extremely premature infants further suggests that ML-guided triage may help close an important gap in access to timely genetic diagnosis.

### Classifier Strengths and Weaknesses

A key strength of our classifier includes a novel target for supervised ML: future genetic evaluation generally (as opposed to use of a specific test or prediction of a diagnostic outcome[21,22]) which, in combination with our center’s robust data warehouse, allowed us to train and test our classifier on a sample an order of magnitude larger than those previously described[22]. Importantly, broadening the target to evaluation by 18 months, rather than evaluation during the NICU stay, produced a more equitable label across GA.

The principal limitation of this work is that our outcome label inherits the biases of historical genetic-evaluation practice. The ideal target would be clinical utility (i.e., the usefulness of genetic testing to one or more stakeholders [29]), but the recent expansion of utility measures for GS and genetic-medicine services in the NICU had not yet occurred when most of our training data accrued[30–33]. Until such measurements accumulate, "did someone choose to evaluate this patient before 18 months of age" remains a defensible proxy that is, by design, broader and more equitable than a target tied to a specific test order or diagnostic outcome.

The training data span 2010-2021 and the calibration and validation 2022-2024, raising the possibility of pandemic-era differences in care that we cannot directly exclude as well as other secular effects related to changes in care phenotypes at our center; however, performance was similar in development and validation cohorts, suggesting the model has not overfit to a specific era of practice [34]. More fundamentally, all data derive from a single Level IV NICU network with a shared EHR, and external validation in NICUs with different EHR vendors, referral patterns, and case-mix is needed before the predicted clinical impact can be expected to generalize. For patients transferred from outside the NCH Neonatal Network, phenotype information from referring centers is incompletely captured **(Supplementary Materials: Cohort and Data),** which may modestly attenuate the day-1 features available to the model for that subgroup.

### Clinical cost and benefit

The HPO-based architecture **(NeoGx)** was selected for its dynamic responsiveness: because HPO terms derive from continuous narrative notes rather than episodic, billing-bound Phecodes, NeoGx updates predictions in real time as documentation accrues, translating into tangible utility across distinct patient subgroups **(Table 2).** Beyond reliably recovering unambiguous phenotypes such as classical aneuploidies, NeoGx flagged over a third of patients whose first genetic evaluation occurred only after NICU discharge. Flagging these infants on Day 1 represents a profound acceleration of care for a vulnerable population that current inpatient practice misses entirely.

NeoGx also recalls most patients who ultimately received GS, regardless of diagnostic yield. Whether this reflects an early phenotypic signature unique to GS-bound patients or simply that such patients accumulate well-developed phenotypes, the clinical implication is the same: NeoGx reliably flags the population most likely to benefit from an rGS-first strategy, replicating expert consensus on easy cases while catching cases current practice reaches late or misses.

Earlier referral alone (Scenario 2) produced only a modest gain, because advancing the referral date does not change the latency of conventional testing pathways. The substantial improvement came when ML triage was paired with rGS ordered upon a positive prediction (Scenario 3), which also outperformed universal rGS without ML triage (Scenario 4, our center’s current standard of care). NeoGx’s contribution is therefore not to replace an rGS-first protocol such as the NSIGHT2 RCT or SeqFirst Neo, but to prioritize infants for front-line genomic testing[35,36].

Because this study is retrospective, we could not assess human-machine interactions once recommendations are made. NeoGx reports daily updates through 28 days of Level IV stay **(Figure 2c-d),** so a deployment could let clinicians act at whatever timepoint best fits their workflow; the limitation is not that the model is locked to day 1, but that we have not measured how clinician behavior interacts with that flexibility. For example, NeoGx recommendations may be similarly accurate across the first several days of a Level IV NICU admission, while clinicians’ willingness to pursue broad genetic testing may increase on the second or third day as other diagnostic studies, such as blood cultures, begin to return. Conversely, without prospective evaluation, we cannot determine how clinicians will respond to negative NeoGx predictions: whether trusted negative results will appropriately reduce low-yield testing, or instead discourage evaluations that clinical judgment would otherwise have prompted.

We also ascribed zero benefit to positive predictions for patients who never received any genetic evaluation, though some may have been evaluated outside our system or simply went unrecognized. More fundamentally, the cost of a false positive depends on the test it triggers: a conventional pathway initiates sequential workup, whereas the rGS-on-flag protocol of Scenario 3 triggers a single rGS, whose results clinicians find useful even when negative, for ruling out genetic etiologies. NeoGx’s false-positive cost is therefore plausibly lower under an rGS-coupled deployment, reinforcing that its value depends on the pathway it is coupled to.

### Future directions and expansions

#### Level III NICUgeneralization

Level III NICUs care for many more patients than Level IV NICUs, but several factors complicate directly extrapolating our approach. The prevalence of genetic illness is higher in Level IV NICUs f∼26% in our training cohort) than would be expected in Level III, where the positive class is likely to be a smaller minority, yielding a more imbalanced learning problem. Level III settings also have a substantially higher proportion of preterm infants[37], the subgroup in which NeoGx already shows reduced recall, which would amplify rather than mitigate that limitation. A direct extension would also need to handle censoring by transfer to Level IV as well as re-curating exclusion criteria for conditions specific to Level III practice. Different labelling and exclusion criteria may enable adaptation of our approach to Level III, but the current NeoGx model should not be deployed in that setting.

#### Phenotyping pipeline portability

Phenotypes for the training data were obtained from clinical text by ClinPhen[38], a rule-based HPO extractor with a fixed lexicon and known limitations around negation, family-history mentions, and abbreviation handling, all errors that propagate as feature noise into NeoGx’s predictions. More fundamentally, clinical-text-based phenotyping is poorly portable across institutions for myriad reasons: note templates, documentation conventions, and even abbreviation usage differ between EHRs. An NLP pipeline tuned at one site is unlikely to perform identically at another without re-engineering. The administrative and computational overhead of obtaining and processing clinical text further hinders portability. Our finding that Phecode-based models achieved strong day-1 performance suggests that structured-code phenotyping is a viable alternative for sites where clinical-text NLP is impractical. However, doing so would come at the cost, as our results show, of the dynamic responsiveness that allowed HPO-based predictions to update with accruing documentation.

#### Prospective validation

The clinical impact reported here was estimated retrospectively, assuming 100% adherence to simulated policies. Real-world CDS never achieves full adherence, and variable clinician response could either amplify the benefit or increase the false-positive cost. A definitive evaluation will therefore require a prospective randomized controlled trial in which the utility of NeoGx’s recommendations is assessed with a validated utility measure[29]. Such an instrument would serve double duty: the same utility measurements that anchor a less label-biased training target would also provide the trial’s endpoint, so both limitations could be resolved through a single measurement infrastructure.

## CONCLUSION

Most neonates with rare genetic disease begin life without a diagnosis, and current practice often delays or misses genetic evaluation in the infants who would benefit most from earlier testing. We addressed this gap by training NeoGx, a machine-learning classifier that predicts future genetic evaluation by 18 months of age, a target broader and more equitable than test-specific or diagnosis-specific outcomes used previously, on the largest such cohort of NICU patients reported to date. In a temporally held-out 2023-2024 validation cohort, NeoGx flagged ∼31% of infants for early genetic workup with a precision of 0.66, and simulated implementation showed that pairing NeoGx-driven referral with rGS on positive predictions raised the share of infants reaching a genetic diagnosis or completed GS within 14 days from 9.5% under current practice to 68.6%, outperforming universal rGS without ML triage. Critically, the absolute benefit was largest for extremely premature infants, the subgroup most underserved by current evaluation pathways, narrowing rather than widening a real-world gap that begins on the first day of NICU admission. If validated prospectively under utility-anchored outcomes, machine-learning triage of this kind has the potential not only to compress the diagnostic odyssey, but to do so most for the infants who today wait longest.

## Supporting information

Supplemental results of bias analysis

## Data Availability

The NeoGx training, evaluation, and analysis code is available as open-source software through GitHub (https://www.github.com/nch-cloud/neogx-iv)[39] and archived in Zenodo (https://doi.org/10.5281/zenodo.20942256)[40]. The repository includes the analysis code used to generate the figures and tables in this manuscript.
The de-identified cohort feature matrices, the trained NeoGx classifier weights (XGBoost .pkl artifacts for HPO and Phecode variants, with and without GA), the corresponding hyperparameter and architecture metadata, and the supporting reference assets (HPO representative-set mappings and PhecodeX dictionary used at inference time) are archived in a separate Zenodo record (https://doi.org/10.5281/zenodo.20615808)[41]. Patient identifiers, dates, and clinical free text were removed prior to release in accordance with the Health Insurance Portability and Accountability Act (HIPAA) Safe Harbor de-identification standard.
Together, these resources are sufficient to reload the trained models and reproduce the headline metrics, subgroup analyses, and simulation results reported in this manuscript.

https://www.github.com/nch-cloud/neogx-iv

https://doi.org/10.5281/zenodo.20942256

https://doi.org/10.5281/zenodo.20615808

## DECLARATIONS

### Ethics approval and consent to participate

The Abigail Wexner Research Institute Institutional Review Board (IRB) approved the study (STUDY00000276) with a waiver of informed consent.

## Consent for publication

Not applicable.

## Availability of data and materials

The NeoGx training, evaluation, and analysis code is available as open-source software through GitHub (https://www.github.com/nch-cloud/neogx-iv)[39] and archived in Zenodo (https://doi.org/10.5281/zenodo.20942256) [40]. The repository includes the analysis code used to generate the figures and tables in this manuscript.

The de-identified cohort feature matrices, the trained NeoGx classifier weights (XGBoost .pkl artifacts for HPO and Phecode variants, with and without GA), the corresponding hyperparameter and architecture metadata, and the supporting reference assets (HPO representative-set mappings and PhecodeX dictionary used at inference time) are archived in a separate Zenodo record (https://doi.org/10.5281/zenodo.20615808) [41], Patient identifiers, dates, and clinical free text were removed prior to release in accordance with the Health Insurance Portability and Accountability Act (HIPAA) Safe Harbor de-identification standard.

Together, these resources are sufficient to reload the trained models and reproduce the headline metrics, subgroup analyses, and simulation results reported in this manuscript.

## Authors’ contributions

Austin A. Antoniou (Conceptualization, Data curation, Formal analysis, Investigation, Methodology, Software, Validation, Writing—original draft, Writing—review & editing), David M. Gordon (Resources, Software, Writing—review & editing), Ashley Kubatko (Project administration, Resources, Supervision, Visualization), Peter White (Conceptualization, Funding acquisition, Resources, Supervision, Visualization, Writing—review & editing), and Bimal P. Chaudhari (Conceptualization, Funding acquisition, Methodology, Project administration, Supervision, Validation, Writing—original draft, Writing—review & editing). All authors reviewed and approved the final manuscript.

## Acknowledgements

We thank the Nationwide Children’s Hospital Foundation Pediatric Innovation Fund for generously supporting this project. The authors would like to acknowledge Rajesh Ganta, Brent Merryman, and Nan Zhang, who obtained data from the Research Data Warehouse and clinical data repositories. Steve Rust and Sven Bambach were consulted on ML study design and approaches to bias evaluation.

The authors used generative AI tools (Anthropic Claude and OpenAI ChatGPT) for editorial assistance (restructuring, polishing, and revision of author-written text) and for engineering and reproducibility tasks (code review, refactoring, regeneration of analysis outputs, and verification that numerical values reported in the manuscript and supplement traced correctly to the analysis notebooks). All scientific concepts, study design, methods, analyses, and final language were developed by the authors; AI-assisted edits and code suggestions were reviewed before adoption, all numerical results were re-verified against the source notebooks, and all citations were independently checked by the team. The authors take full responsibility for the final manuscript, codebase, and reported results.

## Abbreviations

AUC: Area Under the Curve
BW: Birth Weight
CDS: Clinical Decision Support
CI: Confidence Interval
CV: Cross-Validation
Dx: Diagnosis
EHR: Electronic Health Record
GA: Gestational Age
HPO: Human Phenotype Ontology
ICD: International Classification of Diseases
IRB: Institutional Review Board
ML: Machine Learning
NCH: Nationwide Children’s Hospital
NICU: Neonatal Intensive Care Unit
NLP: Natural Language Processing
OR: Odds Ratio
PR: Precision-Recall rGS: rapid Genomic Sequencing
ROC: Receiver Operating Characteristic
SHAP: SHapley Additive explanations

## REFERENCES

1 Marouane A, Olde Keizer R a. CM, Frederix GWJ, et al. Congenital anomalies and genetic disorders in neonates and infants: a single-center observational cohort study. Eur J Pediatr. 2022;181:359–67. doi: 10.1007/s00431-021-04213-w

2 Centers for Disease Control and Prevention (CDC). Improved National Prevalence Estimates for 18 Selected Major Birth Defects—United States, 1999-2001. JAMA. 2006;295:618–20. doi: 10.1001/jama.295.6.618

3 Parker SE, Mai CT, Canfield MA, et al. Updated National Birth Prevalence estimates for selected birth defects in the United States, 2004-2006. Birth Defects Res A Clin Mol Teratol. 2010;88:1008–16. doi: 10.1002/bdra.20735

4 Stallings EB, Isenburg JL, Rutkowski RE, et al. National population-based estimates for major birth defects, 2016-2020. Birth Defects Res. 2024;116:e2301. doi: 10.1002/bdr2.2301

5 Weiner J, Sharma J, Lantos J, et al. How Infants Die in the Neonatal Intensive Care Unit: Trends From 1999 Through 2008. Arch Pediatr Adolesc Med. 2011;165:630–4. doi: 10.1001/archpediatrics.2011.102

6 Simeoni U. How infants die in neonatal intensive care units - a European perspective. Acta Paediatr. 2012;101:552–4. doi: 10.1111/j.1651-2227.2012.02685.x

7 Synnes AR, Berry M, Jones H, et al. Infants with congenital anomalies admitted to neonatal intensive care units. Am JPerinatol. 2004;21:199–207. doi: 10.1055/s-2004-828604

8 Ferreira CR. The burden of rare diseases. Am J Med Genet A. 2019;179:885–92. doi: 10.1002/ajmg.a.61124

9 Haendel M, Vasilevsky N, Unni D, et al. How many rare diseases are there? Nat Rev Drug Discov. 2020;19:77–8. doi: 10.1038/d41573-019-00180-y

10 Messick EA, Backes CH, Jackson K, et al. Morbidity and mortality in neonates with Down Syndrome based on gestational age. J Perinatol. 2023;43:445–51. doi: 10.1038/s41372-022-01514-2

11 Swaggart KA, Swarr DT, Tolusso LK, et al. Making a Genetic Diagnosis in a Level IV Neonatal Intensive Care Unit Population: Who, When, How, and at What Cost? J Pediatr. 2019;213:211–217.e4. doi: 10.1016/j.jpeds.2019.05.054

12 Callahan KP, Clayton EW, Lemke AA, et al. Ethical and Legal Issues Surrounding Genetic Testing in the NICU. NeoReviews. 2024;25:e127–38. doi: 10.1542/neo.25-3-e127

13 Basharat S, Basharat N, Rader T. 2023 Watch List: Top 10 Precision Medicine Technologies and Issues: CADTH Horizon Scan. Ottawa (ON): Canadian Agency for Drugs and Technologies in Health 2023.

14 Petrikin JE, Willig LK, Smith LD, et al. Rapid whole genome sequencing and precision neonatology. Semin Perinatol. 2015;39:623–31. doi: 10.1053/j.semperi.2015.09.009

15 Saunders CJ, Miller NA, Soden SE, et al. Rapid whole-genome sequencing for genetic disease diagnosis in neonatal intensive care units. Sci Transl Med. 2012;4:154ra135. doi: 10.1126/scitranslmed.3004041

16 Smith HS, Ferket BS, Gelb BD, et al. Parent-Reported Clinical Utility of Pediatric Genomic Sequencing. Pediatrics. 2023;152:e2022060318. doi: 10.1542/peds.2022-060318

17 Dimmock D, Caylor S, Waldman B, et al. Project Baby Bear: Rapid precision care incorporating rWGS in 5 California children’s hospitals demonstrates improved clinical outcomes and reduced costs of care. Am J Hum Genet. 2021;108:1231–8. doi: 10.1016/j.ajhg.2021.05.008

18 Maron JL, Kingsmore S, Gelb BD, et al. Rapid Whole-Genomic Sequencing and a Targeted Neonatal Gene Panel in Infants With a Suspected Genetic Disorder. JAMA. 2023;330:161–9. doi: 10.1001/jama.2023.9350

19 Wojcik MH, Del Rosario MC, Agrawal PB. Perspectives of United States neonatologists on genetic testing practices. Genet Med. 2022;24:1372–7. doi: 10.1016/j.gim.2022.02.009

20 Seither K, Thompson W, Suhrie K. A Practical Guide to Whole Genome Sequencing in the NICU. NeoReviews. 2024;25:e139–50. doi: 10.1542/neo.25-3-e139

21 Morley TJ, Han L, Castro VM, et al. Phenotypic signatures in clinical data enable systematic identification of patients for genetic testing. Nat Med. 2021;27:1097–104. doi: 10.1038/s41591-021-01356-z

22 Peterson B, Hernandez EJ, Hobbs C, *et al.* Automated prioritization of sick newborns for whole genome sequencing using clinical natural language processing and machine learning. Genome Med. 2023;15:18. doi: 10.1186/s13073-023-01166-7

23 Juarez EF, Peterson B, Sanford Kobayashi E, et al. A machine learning decision support tool optimizes WGS utilization in a neonatal intensive care unit. NPJ Digit Med. 2025;8:1–4. doi: 10.1038/s41746-025-01458-9

24 Sun X, De Brouwer E, Liu C, et al. Deep learning unlocks the true potential of organ donation after circulatory death with accurate prediction of time-to-death. Sci Rep. 2025;15:13565. doi: 10.1038/s41598-025-95079-7

25 Ramspek CL, Jager KJ, Dekker FW, et al. External validation of prognostic models: what, why, how, when and where? Clin Kidney J. 2020;14:49–58. doi: 10.1093/ckj/sfaa188

26 Lundberg SM, Lee S-I. A Unified Approach to Interpreting Model Predictions. *Advances in Neural Information Processing Systems.* Curran Associates, Inc. 2017.

27 Jagadeesh KA, Birgmeier J, Guturu H, et al. Phrank measures phenotype sets similarity to greatly improve Mendelian diagnostic disease prioritization. Genet Med. 2019;21:464–70. doi: 10.1038/s41436-018-0072-y

28 Janvier A, Leblanc I, Barrington KJ. Nobody likes premies: the relative value of patients’ lives. JPerinatol. 2008;28:821–6. doi: 10.1038/jp.2008.103

29 ACMG Board of Directors. Clinical utility of genetic and genomic services: a position statement of the American College of Medical Genetics and Genomics. Genet Med. 2015;17:505–7. doi: 10.1038/gim.2015.41

30 Pitini E, De Vito C, Marzuillo C, *et al.* How is genetic testing evaluated? A systematic review of the literature. Eur J Hum Genet. 2018;26:605–15. doi: 10.1038/s41431-018-0095-5

31 Smith HS, Rubanovich CK, Robinson JO, et al. Measuring perceived utility of genomic sequencing: Development and validation of the GENEtic Utility (GENE-U) scale for pediatric diagnostic testing. Genet in Med. 2024;26:101146. doi: 10.1016/j.gim.2024.101146

32 Turbitt E, Kohler JN, Brothers KB, et al. The Parent PrU: A measure to assess personal utility of pediatric genomic results. Genet Med. 2024;26:100994. doi: 10.1016/j.gim.2023.100994

33 Hayeems RZ, Luca S, Ungar WJ, et al. The Clinician-reported Genetic testing Utility InDEx (C-GUIDE): Preliminary evidence of validity and reliability. Genet Med. 2022;24:430–8. doi: 10.1016/j.gim.2021.10.005

34 Weishaupt L, Wang T, Schamroth J, et al. Care Phenotypes In Critical Care. 2025;2025.01.24.25320468.

35 Dimmock DP, Clark MM, Gaughran M, et al. An RCT of Rapid Genomic Sequencing among Seriously Ill Infants Results in High Clinical Utility, Changes in Management, and Low Perceived Harm. Am J Hum Genet. 2020;107:942–52. doi: 10.1016/j.ajhg.2020.10.003

36 Wenger TL, Scott A, Kruidenier L, *et al.* SeqFirst: Building equity access to a precise genetic diagnosis in critically ill newborns. The American Journal of Human Genetics. 2025;112:508–22. doi: 10.1016/j.ajhg.2025.02.003

37 Helm BM, Hays M, Swaggart KA, et al. Standardized Criteria for Genomic Testing in the NICU. Pediatrics. 2025;156:e2024069591. doi: 10.1542/peds.2024-069591

38 Deisseroth CA, Birgmeier J, Bodle EE, et al. ClinPhen extracts and prioritizes patient phenotypes directly from medical records to expedite genetic disease diagnosis. Genet Med. 2019;21:1585–93. doi: 10.1038/s41436-018-0381-1

39 Antoniou A. nch-cloud/neogx-iv [software]. GitHub. Accessed June 29, 2026. https://github.com/nch-cloud/neogx-iv

40 Antoniou AA. nch-cloud/neogx-iv: submission [software]. Version 1.0.0. Zenodo. Published June 2026. doi:10.5281/zenodo.20942256

41 Antoniou AA. NeoGx - Predicting Genetic Need in Level IV NICUs: Analysis Dataset [dataset]. Version 1. Zenodo. Published June 2026. doi:10.5281/zenodo.20615809

