## Supplement 1 for "NeoGx: Machine Learning to Predict Genetic Evaluation Need in the Level IV NICU"

### Cohort and Data

#### NCH NICU network and data warehousing

The NCH Neonatal Network consists of 6 Level III NICUs at delivery centers and 1 Level IV NICU in central Ohio. Neonates in NICUs in the Neonatal Networks are cared for by Neonatologists from 3 practices (1 academic, 2 private) and a shared team of Neonatal Nurse Practitioners and Physician Assistants using a shared EHR with clinical and administrative data collected in a single Research Data Warehouse (RDW). Within the neonatal network, patients are assigned a unique identifier which allows self-self linkage of data from delivery centers with data from the Level IV NICU. Patients admitted to the Level IV NICU from outside of the Neonatal Network are not linked to data from the referring center aside from basic demographic and administrative data including birthweight and gestational age at birth. Patients who return to NCH for post-ICU care (including genetics referral/consultation or genetic testing ordered within the NCH EHR) have their data captured in the RDW with self-self linkage to the data from the NICU hospitalization.

#### Data elements

For each subject, birthweight, gestational age at birth (GA), all available lab tests, genetics consults and referrals, and International Classification of Diseases, 9th and 10th Revisions (ICD-9 and ICD-10) for the length of their entire health record were obtained from the NCH RDW and subsequently mapped to Phecodes via the PhecodeX mapping[1]. Clinical note text, from which we computed Human Phenotype Ontology (HPO)-based phenotype features, was collected through the duration of the NICU stay[2].

### Curation

#### Genetic diagnosis codes

The phecodes in the PhecodeX “Genetic” chapter (prefix “GE_”) were labelled according to whether they represented genetic disorders. A subset of 23 “GE_” codes were excluded as not representative of genetic disorders (Table S1). These included codes related to cancer genetics, codes for carrier/trait status for genetic diseases, and diseases which could not manifest or were not relevant to care in the neonatal period (e.g. Thalassemias), or codes which were otherwise underspecified and could not reliably be used to assert the presence of a concrete genetic disorder. All problem list ICD codes mapping under PhecodeX to one of the remaining 285 codes were identified as genetic diagnoses. Classical aneuploidies were identified as any ICD code mapping to a phecode logically entailing GE_160.1 “Trisomies”.

| Phecode | Description | Reason for exclusion |
| --- | --- | --- |
| GE_961.8 | Genetic susceptibility to malignant neoplasm | Cancer |
| GE_961.81 | Genetic susceptibility to malignant neoplasm of breast | Cancer |
| GE_961.82 | Genetic susceptibility to malignant neoplasm of ovary | Cancer |
| GE_961.83 | Genetic susceptibility to malignant neoplasm of prostate | Cancer |
| GE_961.84 | Genetic susceptibility to malignant neoplasm of endometrium | Cancer |
| GE_962.12 | Disorders of tyrosine metabolism | Underspecified |
| GE_962.121 | Tyrosinemia | Underspecified |
| GE_964 | Disorders of carbohydrate metabolism | Underspecified |
| GE_964.1 | Disorders of galactose metabolism | Underspecified |
| GE_967.4 | Hereditary hemochromatosis | Underspecified |
| GE_970 | Hemoglobinopathies | Underspecified |
| GE_970.1 | Sickle-cell disorders | Underspecified |
| GE_970.2 | Thalassemia | Neonatal-irrelevant |
| GE_970.21 | Alpha thalassemia | Neonatal-irrelevant |
| GE_970.22 | Beta thalassemia | Neonatal-irrelevant |
| GE_970.23 | Delta thalassemia | Neonatal-irrelevant |
| GE_970.25 | Thalassemia minor | Neonatal-irrelevant |
| GE_970.4 | Methemoglobinemia | Underspecified |
| GE_982 | Genetic susceptibility of disease, NOS | Underspecified/Carrier |
| GE_983 | Genetic carrier status | Underspecified/Carrier |
| GE_983.1 | Cystic fibrosis carrier | NBS follow-up/Carrier |
| GE_983.2 | Hemoglobin C trait [Sickle-cell trait] | Underspecified/Carrier |

**Table S1. Genetic phecodes excluded from calculation of positive class label**. An ICD code mapping to one of these phecodes, in the absence of any other evidence, was not counted as evidence for inclusion in the positive class for model training. Included here are the phecode, the description provided in the PhecodeX mapping, and our reason for excluding the code. The abbreviation “NBS” refers to newborn screening; because screening for cystic fibrosis is protocolized as a follow-up upon an abnormal newborn screen result, and a CF carrier label generated by such follow-up does not represent a genetic disorder.

#### Genetic lab tests

First, regular expressions were used to match key strings that implied labs were genetic, including: “genome”, “exome”, “sequence”, “chromosome”, “karyotype”. Regular expressions were also applied to find keywords for labs which were decidedly not genetic, such as infectious disease labs, or cancer testing—including cancer genetic/genomic testing, which was viewed as distinct from genetic testing for rare disease diagnosis. Remaining labs with no clear classification were reviewed by Austin A. Antoniou and finally adjudicated by Bimal P. Chaudhari. Subsequently, each lab order was identified as *genetic* or *non-genetic*, with the “genetic” class including ES/GS, single gene or panel next-generation sequencing (NGS) tests, microarrays, karyotypes, and other genetic/epigenetic tests including Fluorescent *In Situ* Hybridization (FISH), methylation analysis, repeat-expansion analysis, and uniparental disomy (UPD) testing. Biochemical tests used to diagnose genetic disorders were also included as qualifying laboratory tests (e.g. dehydrocholesterol level, lysosomal enzyme screen, or oligosaccharide and glycan screen). Lab tests ordered in protocolized fashion upon an abnormal newborn screening result (e.g. sweat chloride test for Cystic Fibrosis) were excluded.

#### Subject-level labels

We identified patients in our study cohort with at least one of the following in the NCH medical record before 18 months of age: (1) an order for a genetic test, (2) a genetic problem list diagnosis code, (3) a consult or referral to a medical geneticist, the underlying principle being: if a patient received genetic evaluation, that was the correct thing to do (independent of whether genetic evaluation led to a diagnosis). Subjects with a genetic evaluation were given an outcome label of *genetic*, which was used as the positive label for ML training. Patients with classical aneuploidies were excluded from training, but were included in calibration and validation cohorts, and were counted among the positive class. The rationale here is that these patients are already typically identified by neonatologists quickly and would likely not benefit from ML assistance, but it is still desirable behavior that the model identifies them with the genetic class.

### Model Development

Static features relevant to NICU hospitalization were calculated, including age at admission to the Level IV NICU, sex, GA at birth, and BW. BW was normalized as a sex- and GA-specific Z-score using the Olsen 2010 intrauterine growth reference (calculated with the PediTools calculator), thereby representing an infant’s size relative to the expected BW distribution for infants of the same sex and GA[3].

Additional features were modeled to represent the evolving phenotype over the first four weeks in the Level IV NICU. For a given day D of a patient’s NICU stay, a subject’s EHR data was truncated to the data accumulated by day D, and a profile was assembled to represent a patient’s phenotype up to that time point. By incrementing D, we capture the gradual evolution of the phenotype over the NICU stay. We considered two different vocabularies to represent phenotypes from EHR data. We mapped International Classification of Diseases (ICD) diagnosis codes to Phecodes using the PhecodeX mapping[1]. We also extracted Human Phenotype Ontology (HPO, version 2025-10-22) terms from unstructured clinical text using the Natural Language Processing (NLP) tool ClinPhen[2,4]. HPO or PhecodeX terms for GA and BW were also removed to avoid correlation with the directly captured GA and BW features (see **Phenotype Features**, below).

A layered search was carried out to determine the optimal phenotype feature representation and classifier-specific hyperparameters. The outer layers enumerated different candidates for feature sets and encodings; accounting for phenotype vocabulary (HPO or Phecodes), the number and specificity of terms used to represent the full vocabulary, and the function used to assign numerical values to a given phenotypic profile (**Supplementary Materials: Phenotype Features**). The inner layers examined architectures and architecture-specific parameters. For each candidate feature set, based on phenotype data available at day 1, the inner search was conducted using 3-fold cross-validation (CV) (see **Classifier and Hyperparameters**, below).

Models were evaluated on the CV fold mean of their Precision-Recall (PR) area under curve (AUC). The best-scoring classifiers using features from each phenotype vocabulary (Phecodes and HPO) and with/without a GA feature were separately analyzed to fully understand the effects of these choices on the quality of predictions. Of particular interest is the question of whether directly encoding GA would improve performance or unduly penalize premature infants, a population expected to have a lower proportion of genetic illness, but in whom symptoms of genetic disorders may be underappreciated or misattributed to prematurity[5].

All four classifier variants were retrained on the whole training cohort, using data available through Level IV NICU day 1, then sigmoid calibrated using the calibration cohort (see **Calibration and Thresholding**, below). After a high-level comparison between the performance of the HPO vs. Phecodes and GA vs. no GA model variants, we focused on the HPO-based model without GA to estimate potential clinical impact.

### Phenotype features

A layered search was used, considering different candidates for which phenotype data source (ICD encounter diagnosis codes vs clinical text) and corresponding phenotype vocabulary (PhecodeX or HPO) was to be used, how the full vocabularies could be compactified to a set of representatives, different formulations for the encoding of feature vector values, architecture, and architecture-specific hyperparameters.

#### Phecode features

All ICD 9- and 10- codes in every subject’s record were mapped to Phecodes using the PhecodeX mapping. Before creating Phecode feature matrices, codes corresponding to the already explicitly encoded GA and BW features or to genetic disorders were excluded from the calculation of features. Exclusions included the following codes.

- Genetic chapter: codes matching the regular expression “GE_.*”
- Small/large for GA: codes matching “NB_850.*” or “NB_851.*”
- GA categories: codes matching “NB_885.*”
- BW categories:codes matching “NB_886.*”
- Retinopathy: codes matching “SO_374.4.*”
  - Retinopathy of prematurity is mapped via SO_374.4 (Retinopathy); it was removed as a potential proxy for prematurity

We represented phenotypes at two levels of specificity; either using “depth 1” or “depth 2” phecodes. Thinking of PhecodeX as a graph, we considered codes of the form “<chapter>_<###>” to be of depth 1, and further specified codes with a single digit after the decimal point to be of depth 2. For example, ID_089 “infections” is depth 1 while ID_089.1 “bacterial infections” is depth 2. After removing genetic, GA, and BW phecodes there were 630 at depth 1 and 2,419 at depth 2.

Each phecode feature was given a binary encoding: 1 if present in a subject’s phenotype profile, otherwise 0. As Phecodes did not have suitable annotations comparable to the Human Phenotype Ontology’s annotations, we did not consider alternate annotation-based encodings.

#### HPO features

The Human Phenotype Ontology (HPO) (2025-10-22 version) was used to computably model phenotypes from electronic health record text. ClinPhen was used to extract HPO terms from any clinical text that was available through the prediction timepoint. We computationally loaded in and navigated the HPO graph using the pyhpo Python package. We start with some definitions of key concepts used to compute feature representations:

- We say an HPO term *x* is a *descendant* of a term *y* (or x *logically entails* y) if there is a path of is-a relationships (directed edges) from *x* to *y* in the directed HPO graph.
  - Equivalently, we can say *y* is an *ancestor* of *x.*
- We say a term *x* is a *child* of a term *y* if there is a single “is-a” relationship (directed edge) from *x* to *y*
  - Equivalently, we can say *y* is a *parent* of *x*
- The set of parents of a term *x*: $\mathrm{pa}\left( x \right)=\left\{ y\in\mathrm{HPO} \right|x is a child of y\}$
- The set of ancestors of a term *x*: $\mathrm{anc}\left( x \right)=\{y\in HPO|x logically entails y\}$
- The set of ancestors of a set *X*: $\mathrm{anc}\left( X \right)=\{y\in HPO|x logically entails y for some x\in X\}$
- The set of descendants of *x*: $\mathrm{desc}\left( x \right)=\{y\in HPO|y logically entails x\}$
- The set of descendants of a set *X*: $\mathrm{desc}\left( X \right)=\{y\in HPO|y logically entails x for some x\in X\}$
- The boundary of a set *X* of terms: $\mathrm{bdy}\left( X \right)=\left\{ x\in X \right|X\cap\mathrm{desc}\left( x \right)=\{x\}\}$ (the terms in *X* with no descendant also belonging to *X*)
- The set of genes associated in HPO annotations to a term *x* or set of terms *X* will be written as $Genes(x)$ or $Genes(X)$
  - The cardinality of the associated gene set will be denoted using $|\cdot|$ notation; e.g. $|\mathrm{Genes}\left( x \right)|$ denotes the cardinality of the gene set of the term *x*.

We borrow the formulation of Information Content (IC) from Phrank, derived from HPO’s phenotype-gene associations, to act as a value function for sets of HPO terms[6]. The IC of a set X of HPO terms is given by the formula

$$IC\left( X \right)=\sum_{x\in\mathrm{anc}(X)} -\log\frac{|\mathrm{Genes}\left( x \right)|}{|\mathrm{Genes}\left( \mathrm{pa}\left( x \right) \right)|}$$

We use this formulation because it allows consistency in calculating IC for individual terms and for sets of terms (letting IC(*x*) = IC({*x*}), and can handle logical redundancy within term sets because of the property IC(*X*) = IC(anc(*X*)).

A total of 993 HPO terms relating to GA, BW, and irrelevant or spuriously tagged phenotypes (e.g. cancer and behavioral terms) were removed from the set of extracted ClinPhen HPO terms before calculation of HPO features:

- HP:0000118 (Phenotypic abnormality)
- HP:0001622 (Premature birth) and all descendants (6 terms)
- HP:0000488 (Retinopathy) and all descendants (18 terms) – proxy for prematurity via retinopathy of prematurity
- HP:0004323 (Abnormality of body weight) and all descendants (30 terms)

Additionally, we excluded cancer-related concepts and behavioral abnormalities. These are phenotype concepts that are unrelated to our predictive task and should not occur at high rates; however, due to idiosyncrasies of our HPO term extraction process, some of these concepts were spuriously tagged in a way we would not from structurally ICD phenotypes. Thus we excluded these terms to avoid the propagation of inappropriate features into the model.

- HP:0002664 (Neoplasm) and all descendants (709 terms) – spurious tagging of this term occurred
- HP:0000708 (Atypical behavior) and all descendants (229 terms)

To compactly represent the full set of over 18,000 concepts in the ontology, we considered strategies for representing the full ontology using subsets of terms; namely graph depth, information potential, semantics/nomenclature, and training data prevalence.

**Depth-based representatives:** One method of specifying a representative subset was to fix a depth – number of steps down from “Phenotypic abnormality” (HP:0000118) in the directed graph. We considered the sets of terms at depths 1, 2, and 3, then removed any logically redundant terms which were entailed by other terms within the representative subset, yielding representative sets of sizes 23, 141, and 716 respectively.

**Information potential-based representatives**: Another means of specifying a set of representative HPO terms leveraged IC. We defined *Information Potential Ratio* (IPR) of a term $x$, to be the fraction

$$IPR\left( x \right)=\frac{IC\left( \mathrm{desc}\left( x \right) \right)}{IC\left( x \right)}$$

Higher IPR indicates a term whose descendants, on aggregate, carry substantially more information than the term itself — a candidate for replacement by its descendants. We considered the representative sets consisting of terms with IPR ≥ 5, IPR ≥ 10, and IPR ≥ 25 (i.e., IC of descendants is at least 5×, 10×, or 25× the IC of the term itself). After calculating boundary term sets to remove logically redundant terms this yielded representative sets of sizes 344, 177, and 82, respectively.

**Semantics-based representatives**: We also defined a set of “named abnormality” representatives using string matching on HPO term names and aliases, designed to capture high-level abnormality terms representing many different systems. Many high level HPO terms of the form “abnormality of X” terms have child terms of the form “abnormal X morphology” and “abnormal X physiology”. The idea was to use morphology, physiology, and other remaining abnormality terms as representatives.

We first selected terms whose preferred name or synonyms contained the strings “abnormal” and “morphology” to find all morphological abnormalities to form X_morph_. Similarly, we found all terms whose labels or synonyms contained “abnormal” and “physiolog” to form X_phys_. For the remaining abnormalities which couldn’t be classified as either morphological or physiological abnormalities, we found terms with “abnormal” but neither “morpholog” or “physiolog” in their label or synonyms to form X_other_. To avoid redundancy, we removed all descendants of X_morph_ and X_phys_ from X_other_, then kept only the most specific non-redundant terms from each. That is, the final set of named-abnormality representatives was

$$\mathrm{bdy}\left( X_{morph} \right)\cup\mathrm{bdy}(X_{phys})\cup\mathrm{bdy}(X_{other}\setminus\mathrm{desc}\left( X_{morph} \right)\setminus\mathrm{desc}\left( X_{phys} \right))$$

This semantically defined set of representatives had 740 terms.

**Prevalence-based representatives**: We determined empirically which terms might be useful representatives by measuring the prevalence of all HPO terms in the development set of patients with birth date 2021 or earlier. For each individual, we propagated their observed HPO terms logically upward through the HPO graph, so that the presence of any term also counted toward the prevalence of any terms that it logically entailed (e.g. if “small head” was present in an individual’s phenotypic profile, then we inferred that so too were “abnormal skull size”, “abnormality of head or neck”, and all other ancestor terms of “small head”). We considered as representatives the *saturated* and *boundary* sets of terms at a range of prevalence thresholds, where “saturated” refers to retaining *all* terms at a prevalence threshold and “boundary” refers to keeping only the most specific, non-redundant terms (Table S2). The set of 2,535 saturated representatives at prevalence 0.001 was ultimately chosen for the main classifier NeoGx.

| **Prevalence threshold** | **# representatives (saturated)** | **# representatives (boundary)** |
| --- | --- | --- |
| 0.1 | 291 | 106 |
| 0.05 | 464 | 176 |
| 0.02 | 749 | 312 |
| 0.01 | 1,073 | 464 |
| 0.005 | 1,442 | 644 |
| 0.001 | 2,535 | 1,253 |

**Table S2. Size of HPO prevalence-based representative sets.** Using a range of thresholds for inclusion in the feature set based on prevalence in our training data, we see the number of all HPO terms meeting the prevalence threshold including logically entailed terms (saturated set of representatives) and including only the most specific term meeting the threshold (boundary set, in which there are no ancestor-descendant pairs).

#### HPO feature encodings

Letting $X_{subj}$ be the set of HPO terms in a subject’s EHR text and letting $x$ be one of the representative feature terms, several formulations of feature encoding were considered. First, binary encoding indicated whether *any* concept logically entailing $x$ was present in the subject’s phenotype profile; that is:

$$\mathrm{binary}_{x}\left( \mathrm{subject} \right)=\left\{ \begin{aligned} 1 \mathrm{if} X_{subj}\cap\mathrm{desc}\left( x \right)\neq\emptyset\\ 0 \mathrm{if}X_{subj}\cap\mathrm{desc}\left( x \right)=\emptyset\end{aligned} \right.$$

Secondly, a *lower IC* encoding measured the total IC of all descendants of $x$ in the subject’s phenotype profile:

$$IC_{x}^{\mathrm{lower}}\left( \mathrm{subject} \right)=\mathrm{IC}[X_{subj}\cap\mathrm{desc}\left( x \right)]$$

Finally, a *bidirectional IC* encoding measured the IC of any logical descendant *or* *ancestor* terms present in the subject’s phenotype profile:

$$IC_{x}^{\mathrm{bidirectional}}\left( \mathrm{subject} \right)=\mathrm{IC}[X_{subj}\cap(\mathrm{desc}\left( x \right)\cup\mathrm{anc}\left( x \right))]$$

The bidirectional IC approach passes on logical entailment relationships between phenotype terms to the classifier, affording the model the opportunity to learn parts of the HPO’s hierarchy.

### Classifier and Hyperparameters

#### Classifiers

NeoGx was implemented as a pipeline in Python scikit-learn (version 1.7.2) using the sklearn.pipeline.Pipeline class, consisting of a scaler (the sklearn.preprocessing.StandardScaler class) and a classifier object, implemented using scikit-learn objects when possible, except for the XGBoost classifier implemented using the Python xgboost package (version 2.1.4). The architectures considered were Naïve Bayes (sklearn.naive_bayes.GaussianNB), Logistic Regression (sklearn.linear_model.LogisticRegression), Random Forest (sklearn.ensemble.RandomForestClassifier), and Gradient Boosted Trees (xgboost.XGBoostClassifier). These architectures were chosen for portability and interpretability, as well as their suitability for data at the size and sparsity of ours. Deep learning approaches were judged to require a larger data set than ours.

#### Hyperparameters and randomized search

In addition to different HPO representative feature sets and encodings of those features, we considered several ML architectures and corresponding hyperparameter combinations by sampling parameter spaces using sklearn’s RandomizedSearchCV function. Model performance was ranked by fold mean CV Precision-Recall (PR) Area Under the Curve (AUC) score, using sklearn.metrics.average_precision_score. For reproducibility, we set random_state=614.

Hyperparameter spaces defined for each function are outlined below, naming the parameter and the distribution of values sampled.

Sampling distribution functions:

- scipy.stats.randint
- scipy.stats.uniform
- scipy.stats.loguniform

Naïve Bayes (5 iterations for randomized search):

space_gnb = {

"clf": [clf_name_to_init['GaussianNB']],

"clf__var_smoothing": loguniform(1e-12, 1e-6),

}

Logistic Regression (15 iterations for randomized search):

space_lr = {

"clf": [clf_name_to_init['LogisticRegression']],

"clf__class_weight": [None, 'balanced'],

"clf__solver": ["saga"],

"clf__penalty": ["l1", "l2", None],

"clf__C": loguniform(1e-3, 1e2), # [0.001, 100]

"clf__fit_intercept": [True, False],

}

Random Forest (30 iterations for randomized search):

space_rf = {

"clf": [clf_name_to_init['RandomForestClassifier']],

"clf__n_estimators": randint(200, 1001), # 200–1000

"clf__max_depth": [None] + list(randint(5, 51).rvs(10)), # mix None (option for no max) + sampled depths

"clf__min_samples_split": randint(2, 21),

"clf__min_samples_leaf": randint(1, 11),

"clf__max_features": ["sqrt", "log2", 0.3, 0.5, 0.8],

"clf__bootstrap": [True],

"clf__class_weight": [None, 'balanced_subsample']

}

XGBoost (50 iterations for randomized search):

space_xgb = {

"clf": [clf_name_to_init['XGBClassifier']],

"clf__n_estimators": randint(300, 1001),

"clf__learning_rate": loguniform(1e-2, 3e-1), # [0.01, 0.3]

"clf__max_depth": randint(3, 10),

"clf__min_child_weight": randint(1, 11),

"clf__subsample": uniform(loc=0.5, scale=0.5), # [0.5, 1.0]

"clf__colsample_bytree": uniform(loc=0.5, scale=0.5), # [0.5, 1.0]

"clf__gamma": [0.0, 0.1, 0.3, 1.0],

"clf__reg_alpha": loguniform(1e-4, 10), # [0.0001, 10]

"clf__reg_lambda": loguniform(1e-1, 10),

"clf__scale_pos_weight": [1, spw],

}

(spw = max(1.0, n_neg / max(1, n_pos)), where n_neg and n_pos are the counts of class-0 and class-1 in the training cohort)

### Selected Parameters

Here we enumerate the parameters for the models highlighted in the Results; we chose the Phecode and HPO-based models with best 3-fold mean PR AUC score, and also evaluated those models with GA feature removed.

- Phecode-based model
  - Representatives: phecode depth 2 (2,419 phecode features + 4 static)
    - Phecode+GA model: 2,423 total features
    - Phecode no-GA model: 2,422 features
  - Encoding: binary
  - Classifier: XGBoost
  - Hyperparameters:
    - max_depth = 3
    - n_estimators = 771
    - colsample_bytree = 0.756
    - gamma = 0.1
    - learning_rate = 0.047
    - min_child_weight = 2
    - reg_alpha = 0.0003
    - reg_lambda = 0.581
    - scale_pos_weight = 1
    - subsample = 0.745
- HPO-based model
  - Representatives: saturated prevalence 0.001 (2,535 HPO features + 4 static) (**Table S2**)
    - HPO+GA model: 2,539 total features
    - HPO no-GA model: 2,538 features
  - Encoding: bidirectional IC
  - Classifier: XGBoost
  - Hyperparameters:
    - max_depth = 6
    - n_estimators = 419
    - colsample_bytree = 0.652
    - gamma = 0.3
    - learning_rate = 0.028
    - min_child_weight = 2
    - reg_alpha = 0.092
    - reg_lambda = 2.864
    - scale_pos_weight = 1
    - subsample = 0.687

### Calibration and Thresholding

The subjects in the calibration cohort (N=1,080) were held out from training and used only for probability calibration and setting a decision threshold for dichotomized model predictions and subsequent metrics.

#### Probability Calibration

After all classifier variants were trained on the full training set (N=11,201) using phenotype data available at NICU day 1, they were calibrated using sklearn’s CalibratedClassifierCV on the day-1 features for the calibration cohort (N=1,080) (**Figure S1**).

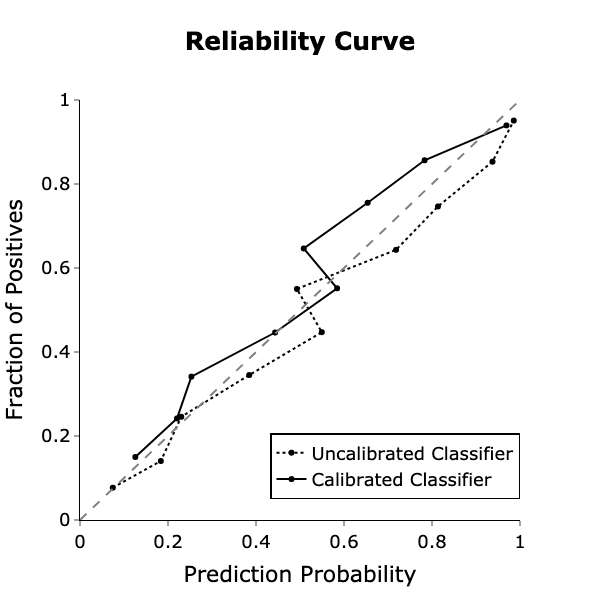

**Figure S1. Classifier calibration curve**. The dotted black curve shows, at each decile, the relationship between NeoGx’s prediction probability score range (expected proportion of positives) and the true fraction of positives in the validation cohort. The solid black curve shows the same relationship after sigmoid calibrating NeoGx. The dashed line indicates the calibration curve of a perfectly calibrated classifier.

#### Decision Boundary

A per-patient prediction probability was computed as the maximum, across NICU-stay days 1-7, 14, 21, and 28 of the calibrated model's predicted probability built from EHR data accumulated through day D. Probability scores for days after a patient's discharge from the Level IV NICU were set to zero so that the per-patient maximum reflects only the time the patient was actually under Level IV care. The clinical operating threshold was set at the 70th percentile of these per-patient maximum scores within the calibration cohort (N = 1,080). For the NeoGx (HPO-without-GA) classifier this yielded a probability threshold of 0.487; for the alternative HPO+GA, Phecode-without-GA, and Phecode+GA variants the corresponding thresholds were 0.476, 0.298, and 0.304, respectively. All downstream binary classification metrics (precision, recall, positive-prediction rate, and per-subgroup analyses) use this threshold to convert calibrated probabilities into positive/negative recommendations.

### Bias Evaluation

Bias in this problem may stem from the underlying representation of demographic groups in our data, genetics utilization practices to date and resulting consequences on labelling, or the predictive behavior of the model itself[7]. Biases related to the model’s predictive behavior were assessed by a similar comparison between each subpopulation’s confidence intervals for the proportion of positive ML predictions, precision, and recall. We considered the impact of these factors across subpopulations defined by sex, race, ethnicity, and GA[8].

#### Subgroup definitions

To account for extremely small demographic subgroups in our cohort, we merged some of the racial and ethnic categories. Racial groups present in at least 5% of the full cohort were left as is; these were “White”, “Black or African American”, and “Multiple”. The remaining groups, “Asian”, “Native American or Alaska Native”, and “Native Hawaiian or Pacific Islander” which each made up less than 5% of the cohort were collected into a single “Other/Unknown” category. Though “Unknown” met the 5% threshold, it is not a substantive category on its own so we merged it with “Other” for statistical comparisons (pre-merge counts of race are still available in **Table 1**). Ethnicity was re-categorized into “Hispanic or Latino”, “Not Hispanic or Latino”, and all other values were collected into an “Other/Unknown” category.

Gestational age was discretized into 3 bins:

- “≥ 35 weeks”
- “Premature”, with GA in the interval [28, 35); that is, 28 ≤ GA < 35
- “Extremely premature”, with GA in [22,28); that is, 22 ≤ GA < 28

Our motivation for the 35 week threshold was to match NCH Neonatal Network protocol, under which babies born at GA < 35 weeks are automatically admitted to the NICU.

#### Regression framework

Logistic regressions with subgroup as covariate were carried out to measure the differences in positive class prevalence and model performance via odds ratios derived from the regression coefficients. See the logistic regression formulae below for details:

- *Positive class prevalence*: positive_class ~ attribute
  - Calculated on full validation cohort
  - Variants of this regression were carried out for variants of the target outcome restricted to shorter time windows: genetic evaluation
    - by Level IV admission;
    - by NICU day 7;
    - by NICU day 14;
    - by NICU discharge;
    - by age 18 months (the chosen definition for the positive class)
- *Positive prediction rate*: positive_prediction ~ attribute
  - Calculated on full validation cohort
  - positive_prediction is a function of model variant (HPO or Phecode; GA or no GA)
- *Recall*: positive_prediction ~ attribute
  - calculation restricted to positive class
  - positive_prediction is a function of model variant (HPO or Phecode; GA or no GA)
- *Precision*: positive_class ~ attribute
  - Calculation restricted to those predicted positive, dependent on model variant considered

These were carried out for attributes including Sex, Race, Ethnicity, and GA (binned as described above) and for each classifier variant: HPO/Phecode based, including or excluding explicit GA feature. For odds ratio calculations, the majority subgroup with respect to each attribute was set as the reference group. For comparisons across sex, the reference subgroup was “Male”; for race, the reference was “White”; for ethnicity, the reference was “Not Hispanic or Latino”; and for GA, the reference was “GA ≥ 35”. For example, odds ratios for GA subgroups are relative to the GA ≥ 35 subgroup.

#### Bias in positive class prevalence

There was no statistically significant difference in the prevalence of the positive class between subgroups defined by sex, race, or ethnicity, even considering all follow-up windows enumerated above (by Level IV admission; NICU day 7; NICU day 14; NICU discharge; or 18 months). However, premature infants had a lower rate of genetic utilization. In the first week of NICU admission, infants born at <28 weeks GA were evaluated at roughly one-third the rate of term infants (OR 0.35, 95% CI 0.24–0.53), and infants born at 28–35 weeks at roughly half (OR 0.55, 95% CI 0.42–0.73). By 18 months, these gaps had narrowed substantially (GA<28: OR 0.70, p = 0.021; GA 28–35: OR 0.72, p = 0.012).

#### Bias in model performance

No differences were found across sex, race, or ethnicity subgroups with respect to any of the performance metrics measured: genetic utilization, positive prediction rate, precision, and recall. Performance differences were apparent with respect to GA. NeoGx showed weaker performance in extremely premature (GA < 28 weeks) subgroup than in the babies born at GA ≥ 35 weeks; the positive prediction rate was lower for extremely premature babies (OR = 0.710, CI 0.528 – 0.954, p = 0.043), as were the precision (OR = 0.314, CI 0.188 – 0.527, p < 0.001) and recall (OR = 0.314, CI 0.187 – 0.528, p < 0.001). Notably, however, these differences were milder than their counterparts in the HPO with GA model (**Figure S2**). A comprehensive table of subgroup comparisons for all model variants appears in **Supplementary Results – Bias Tests**.

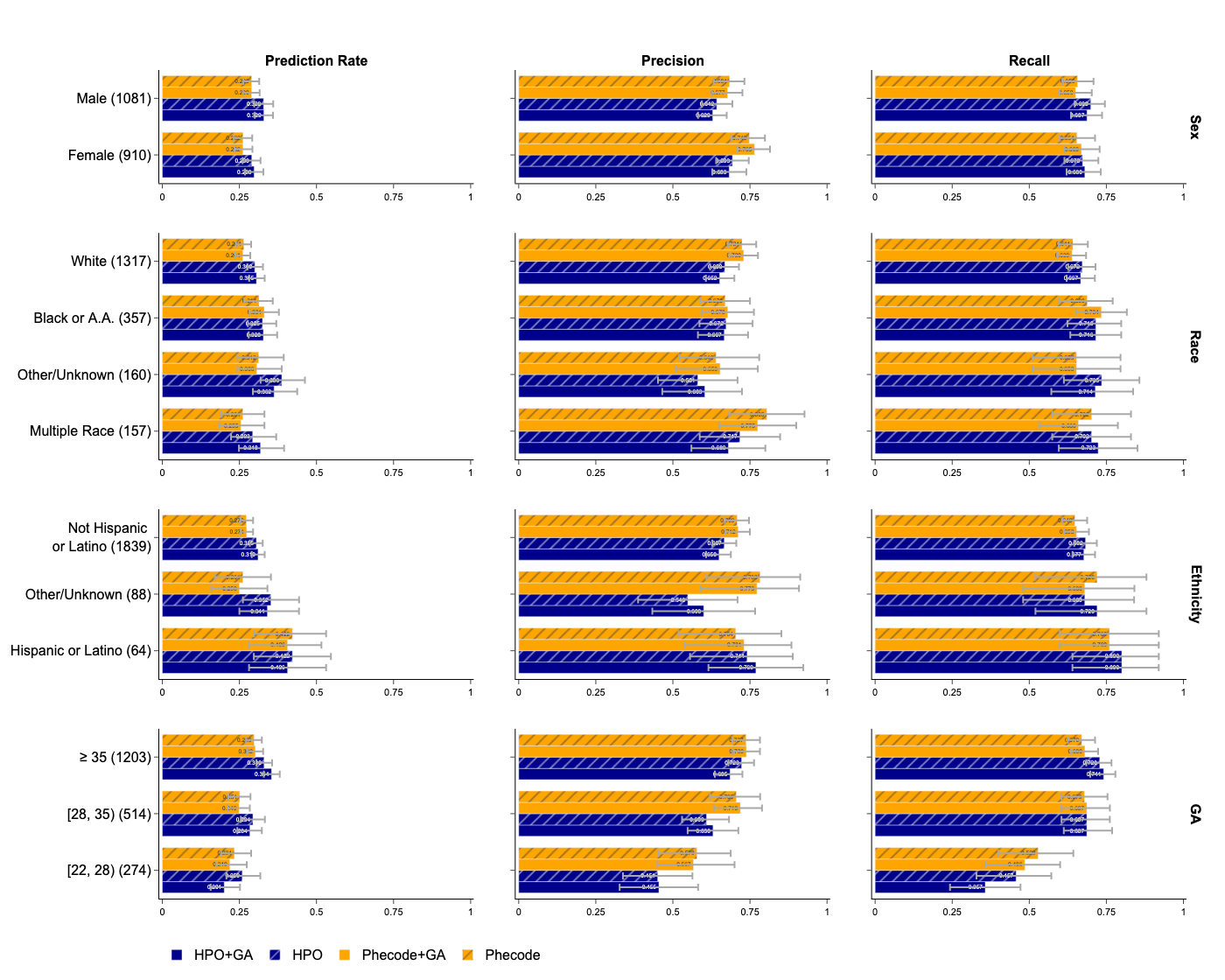

**Figure S2. ML bias by patient characteristics.** Shown above are 16 subplots; rows correspond to population characteristic: sex, race, ethnicity, gestational age. Columns correspond to quantity measured: fraction of the population with genetic testing utilization within 18 months, rate of positive ML prediction, precision, and recall. Each subplot (e.g. the top rightmost set of bars) shows the given metric (e.g. recall) for each subpopulation corresponding to the given patient characteristic (e.g. sex). Error bars indicate a 95% confidence interval for the given metric, and the black dashed lines show the value of each metric for the full validation cohort.

Recall was lower in preterm infants, an effect with two plausible contributors: phenotype features available at NICU day 1 are less informative when the eventual genetic phenotype has not yet manifested, and historical evaluation in this group was systematically delayed relative to term infants, making it harder to predict early an event the real world produces late. Whether this lower recall reflects a true difference in clinical need or a learned bias against preterm infants is not directly resolvable from our data, but recent evidence for the utility of genetic testing in conditions like cerebral palsy, where prematurity is a risk factor and genetic etiologies are well-described, supports bias as a substantial contributor[31–35]. Reassuringly, the most informative features per SHAP values had face validity. We observed that explicit gestational-age encoding shifted both HPO and Phecode models away from positive predictions in preterm infants, a learned bias that we identified and addressed by selecting the GA-free HPO variant as the final NeoGx model. Even at its lower positive-prediction rate in this group, simulated NeoGx intervention recommended genetic evaluation for more <28-week infants than current practice and produced a larger absolute reduction in age at first evaluation (33 days vs. 15 days cohort-wide), narrowing rather than widening the GA gap.

### Feature Importance

The main body showed features with top mean(|SHAP|) scores for the HPO model with no GA feature, the main model considered for the manuscript. That table aggregated calculated SHAP values over all observations in the validation cohort; however, SHAP allows for explanations at the individual prediction level. This can be visualized in a beeswarm plot (**Figure S3**).

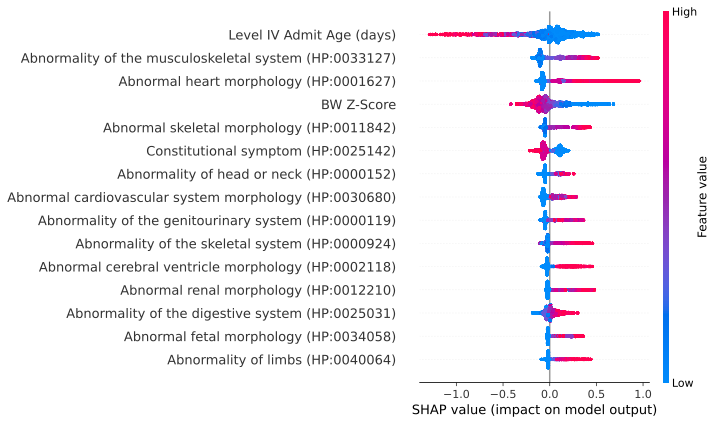

**Figure S3.** SHAP Beeswarm plot. A visualization from the SHAP package of feature importance. The features of NeoGx with highest mean absolute Shapley values are shown on the vertical axis, and the horizontal axis shows the SHAP value, indicating the degree to which the feature influences positive predictions. Individual points correspond to individual subjects in the validation cohort, and the color of the point indicates whether the input value for the given feature was high or low. Abbreviations: “BW”: “Birth weight”.

By analogy with Table 3, we include summaries of feature importance for the alternate models: HPO with GA, and Phecode models with and without GA (**Table S3**).

| **Feature Name** | **Mean Absolute SHAP Value** | **Mean SHAP Value** | **SHAP-Feature Spearman Correlation** | **Inferred Direction** |
| --- | --- | --- | --- | --- |
| **Model: HPO with GA** | | | | |
| Level IV Admit Age (days) | 0.17 | -0.03 | -0.84 | - |
| Abn. musculoskeletal system (HP:0033127) | 0.14 | 0.03 | 0.85 | + |
| Abnormal heart morphology (HP:0001627) | 0.13 | 0.03 | 0.82 | + |
| BW Z-Score | 0.11 | -0.01 | -0.93 | - |
| GA (weeks) | 0.11 | -0.01 | 0.67 | + |
| Abnormal skeletal morphology (HP:0011842) | 0.09 | 0.02 | 0.72 | + |
| Abn. cardiovascular morphology (HP:0030680) | 0.08 | 0.01 | 0.84 | + |
| Abnormality of the skeletal system (HP:0000924) | 0.08 | 0.01 | 0.08 | + |
| Constitutional symptom (HP:0025142) | 0.07 | 0.00 | -0.89 | - |
| Abnormality of head or neck (HP:0000152) | 0.07 | 0.02 | 0.87 | + |
| **Model: Phecode without GA** | | | | |
| Congenital anomalies of the heart (CM_763) | 0.23 | -0.04 | 0.62 | + |
| Level IV Admit Age (days) | 0.18 | -0.02 | -0.87 | - |
| CM of nervous system (CM_750) | 0.17 | -0.03 | 0.42 | + |
| BW Z-Score | 0.14 | -0.01 | -0.72 | - |
| Multiple congenital malformations (CM_776) | 0.12 | 0.01 | 0.29 | + |
| Food+fluid intake signs/symptoms (SS_815) | 0.10 | 0.00 | 0.74 | + |
| CM of tongue, mouth and pharynx (CM_754) | 0.09 | 0.00 | 0.27 | + |
| Congenital deformities of skull/face/jaw (CM_755) | 0.08 | -0.01 | 0.32 | + |
| Nox. substances via placenta/breast milk (NB_879) | 0.07 | 0.00 | -0.50 | - |
| Respiratory distress of newborn (NB_854.1) | 0.06 | -0.01 | -0.81 | - |
| **Model: Phecode with GA** | | | | |
| Congenital anomalies of the heart (CM_763) | 0.23 | -0.04 | 0.62 | + |
| GA (weeks) | 0.19 | 0.00 | 0.66 | + |
| CM of nervous system (CM_750) | 0.16 | -0.03 | 0.43 | + |
| BW Z-Score | 0.15 | -0.01 | -0.71 | - |
| Level IV Admit Age (days) | 0.13 | -0.01 | -0.86 | - |
| Multiple congenital malformations (CM_776) | 0.12 | 0.01 | 0.28 | + |
| Food+fluid intake signs/symptoms (SS_815) | 0.10 | 0.00 | 0.74 | + |
| CM of tongue, mouth and pharynx (CM_754) | 0.09 | 0.00 | 0.26 | + |
| Cong. deformities of skull, face, and jaw (CM_755) | 0.08 | -0.01 | 0.29 | + |
| Nox. substances via placenta/breast milk (NB_879) | 0.08 | 0.00 | -0.49 | - |

**Table S3. Feature importance for alternate models**. By analogy with Table 3 in the main text, this table shows the top 10 features, ranked by highest absolute mean SHAP score, for each of the alternate models (HPO with GA, Phecode with GA, Phecode without GA). Mean absolute SHAP score, taken over the validation cohort for each feature, indicates the relative importance of a feature. Mean SHAP score, is one measure for indicating directionality of the importance, with positive indicating that high feature values lead to higher prediction scores. The SHAP-feature value spearman correlation is presented as an alternate measure of directionality, and we use the sign of this correlation as the inferred direction of each feature’s importance. Some HPO/Phecode labels have been abbreviated to fit on a single line, but the term IDs are included to dispel any ambiguity. In particular, the Phecode “Newborn affected by noxious substances transmitted via placenta or breast milk” is shortened to “Nox. substances via placenta/breast milk”. Abbreviations: “Abn”: “Abnormal”; “Cong.”: “Congenital”; “CM”: “Congenital Malformation”; “GA”: “gestational age”.

Notably, GA was ranked as the second-most-important feature in the alternate Phecode+GA model and fifth in the HPO+GA model, consistent with our finding that GA encoding shifted predictions away from the most premature infants and further motivating our choice of the HPO-without-GA variant as the final NeoGx model.

### Estimated clinical cost and benefit

#### Simulated Intervention Scenario Definitions

For subjects who received genetic evaluation, their *first evaluation age* is defined as their age in days at first *genetic evaluation event*: genetic lab test, consult, referral, or diagnosis code. Their *evaluation duration* is defined as the number of days between their first evaluation event and the most recent genetic evaluation event in their health record. We refer to the sum of a subject’s first evaluation age and duration as their *last evaluation age*. We simulated interventions which could affect (i) the *first evaluation age* by using ML predictions to direct clinicians to begin genetic testing earlier and (ii) the *evaluation duration* by making rGS the first-line test.

To model the ML intervention, we calculated *ML start age* by replacing actual first evaluation age with the first day on which a patient received a positive ML prediction; if they never received a positive prediction, their start age remained unchanged. To model the rGS intervention, we assume rGS is the first test ordered and that no subsequent testing is needed. Thus, the *rGS duration* is sampled from a Poisson random variable with lambda parameter equal to 7 to approximate the reported turnaround time (in days) of rGS testing[9]. If the actual testing duration was shorter, it was used instead, under the assumption that rGS would not hinder providers’ speed in concluding genetic evaluation.

We then considered 5 scenarios (including baseline of actual care) for applying combinations of these interventions:

- Scenario 1 – “Actual”: use actual evaluation start age and duration.
- Scenario 2 – “ML”: use ML start age, make no change to testing duration.
- Scenario 3 – “ML + rGS (limited)”: use ML start age and rGS duration *for ML predicted individuals only*. Others keep their actual evaluation duration time.
- Scenario 4 – “rGS (all)”: do not change start age and use rGS duration for *everybody*.
- Scenario 5 – “ML + rGS (all)”: use ML start age and rGS duration for *everybody*.

To evaluate the impact of ML on genetic evaluation start age, we compared the mean start ages for scenarios 1 and 2. To estimate impact of the combined effects of ML and rGS, we compared across scenarios the mean age at last genetic evaluation and the percentage of positively labelled subjects with a diagnosis or negative GS by Level IV NICU day 14.

#### Reduction of Time to Initial Testing with ML Recommendations

We additionally examined subpopulation differences in real genetic eval age (Scenario 1) and ML-aided start (Scenario 2). There was no apparent difference across subgroups defined by sex, race, or ethnicity; however, we see that the expected improvement in first genetic evaluation age increased in descending order of binned GA (**Figure S4**).

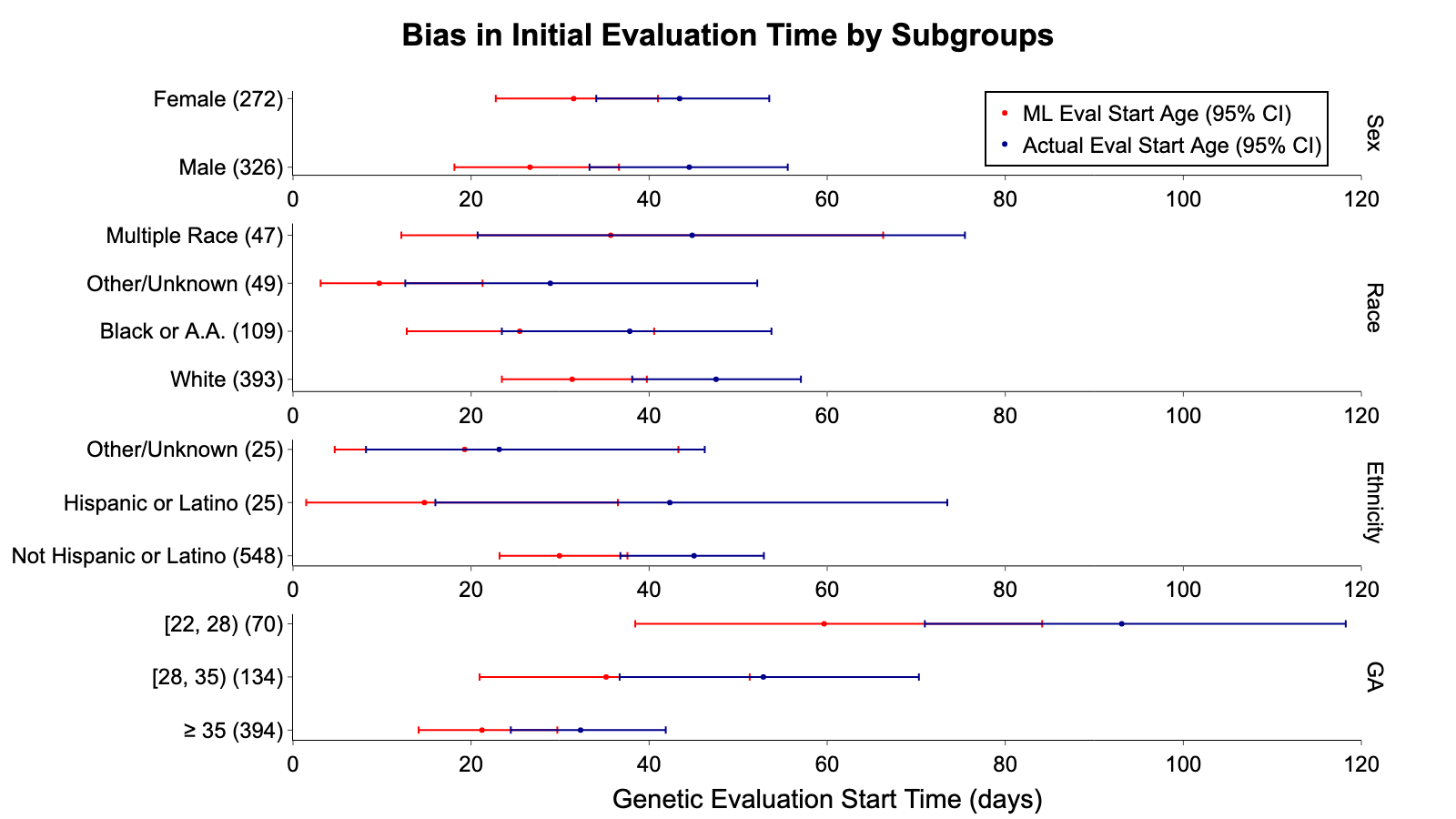

**Figure S4. Test initiation time, with and without ML, by patient characteristics.** For the genetic subset of the validation cohort, time to initial genetic utilization is defined and measured in days. Each plot splits the validation cohort into subgroups defined by the given patient characteristic: sex, race, ethnicity, GA. The vertical axis for each subplot shows each subgroup of the positive class with parenthetical counts of the numbers of members. The horizontal axis measures the time to initial genetic evaluation, in days. We see the mean and 95% confidence interval shown both without ML intervention (Scenario 1) in blue and with ML intervention (Scenario 2) in red.

#### Simulated time to genetic odyssey conclusion

Here we measure the accrual of odyssey completion over time when simulating Scenarios 1-5 as defined in the Methods. We focus on the clinically relevant outcome of whether, under a given scenario, each patient received a genetic diagnosis or GS (Dx/GS for brevity) within 14 days. Compared to real genetic testing history (Scenario 1), in which only 9.5% of actual cases received Dx/GS by age 14 days, initiating testing based on ML recommendations (but not changing any of the test selection; ML-Aided Start (Scenario 2) increased this to 11.9%. Enacting ML+rGS (limited) (Scenario 3) led to 14-day Dx/GS in 68.6% of cases (p<10^-87^ vs. Scenario 2, Z-test). Under rGS (All) (Scenario 4), only 55.2% received Dx/GS (p<10^-5^ vs Scenario 3, Z-test). Finally, an “optimal” policy where the only genetic test ordered is rGS and testing initiation is informed by ML predictions (Scenario 5) brought 14-day Dx/GS to 75.9% (p<10^-12^ vs Scenario 4, Z-test).

A comparison of the Kaplan-Meier curves by log rank test found no statistically significant difference between Scenarios 1 and 2. Scenario 3, 4, and 5 are all superior to Scenario 2 (p<10^-22^). Scenario 3 is bested by Scenario 4 (rGS for all, without ML, p=0.045) and by Scenario 5 (rGS for all, with ML, p=0.003). Most notably, Scenario 5 improves on Scenario 4 (p<10^-8^), which matches the current NCH standard of care, simply by adding the ML intervention to accelerate time to initial genetic evaluation (**Figure S5**).

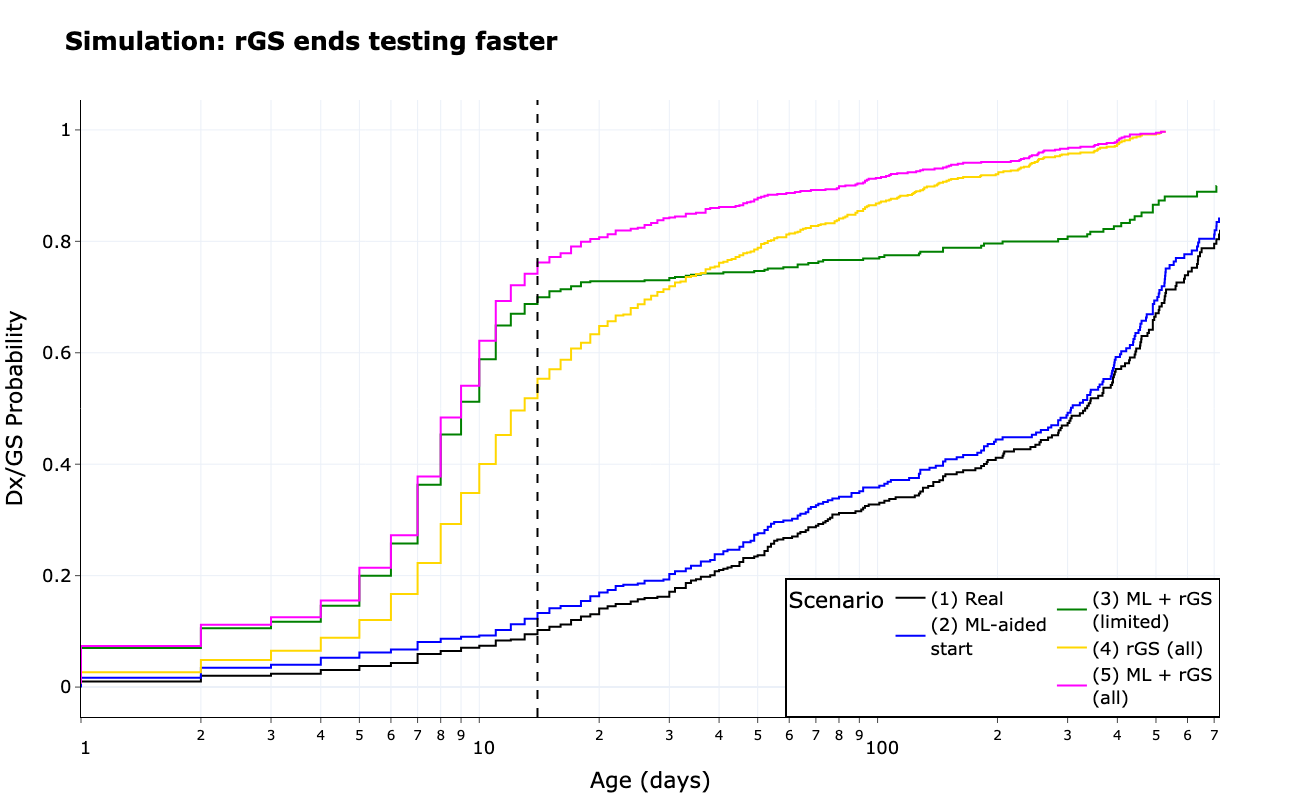

**Figure S5. Time to genetic odyssey completion under simulated recommendation policies.** Under each scenario (see Methods) we measure the time taken for all positively labelled subject in the test set to complete their genetic odyssey. The horizontal axis measures the age in days, log-10 scaled. The vertical axis measures the fraction of all positively labelled subjects in the test set – those with known genetic testing or diagnoses – who have received a genetic diagnosis or GS test by the specified timepoint. The different policies include: *Real* (black); *ML-aided start* (blue) doesn’t change the tests ordered, just the time at which testing starts; *ML + rGS (limited)* (orange) assumes rGS is ordered at the time a positive prediction is made; *rGS (all)* (purple) replaces all initial test orders with rGS but does not change the time at which the test was ordered; *only ML + rGS (all)* (pink) replaces all initial orders with rGS, and accelerates the initial order time in the case of positive predictions.

### References for Supplementary Materials

1	Shuey MM, Stead WW, Aka I, *et al.* Next-generation phenotyping: introducing phecodeX for enhanced discovery research in medical phenomics. *Bioinformatics*. 2023;39:btad655. doi: 10.1093/bioinformatics/btad655

2	Gargano MA, Matentzoglu N, Coleman B, *et al.* The Human Phenotype Ontology in 2024: phenotypes around the world. *Nucleic Acids Res*. 2024;52:D1333–46. doi: 10.1093/nar/gkad1005

3	Chou JH, Roumiantsev S, Singh R. PediTools Electronic Growth Chart Calculators: Applications in Clinical Care, Research, and Quality Improvement. *J Med Internet Res*. 2020;22:e16204. doi: 10.2196/16204

4	Deisseroth CA, Birgmeier J, Bodle EE, *et al.* ClinPhen extracts and prioritizes patient phenotypes directly from medical records to expedite genetic disease diagnosis. *Genet Med*. 2019;21:1585–93. doi: 10.1038/s41436-018-0381-1

5	Messick EA, Backes CH, Jackson K, *et al.* Morbidity and mortality in neonates with Down Syndrome based on gestational age. *J Perinatol*. 2023;43:445–51. doi: 10.1038/s41372-022-01514-2

6	Jagadeesh KA, Birgmeier J, Guturu H, *et al.* Phrank measures phenotype sets similarity to greatly improve Mendelian diagnostic disease prioritization. *Genet Med*. 2019;21:464–70. doi: 10.1038/s41436-018-0072-y

7	Mehrabi N, Morstatter F, Saxena N, *et al.* A Survey on Bias and Fairness in Machine Learning. *ACM Comput Surv*. 2021;54:115:1-115:35. doi: 10.1145/3457607

8	Callahan KP, Clayton EW, Lemke AA, *et al.* Ethical and Legal Issues Surrounding Genetic Testing in the NICU. *NeoReviews*. 2024;25:e127–38. doi: 10.1542/neo.25-3-e127

9	Marom D, Mory A, Reytan-Miron S, *et al.* National Rapid Genome Sequencing in Neonatal Intensive Care. *JAMA Netw Open*. 2024;7:e240146. doi: 10.1001/jamanetworkopen.2024.0146
